# Does hospital overload increase the risk of death when infected by SARS-CoV-2?

**DOI:** 10.1101/2024.08.26.24312569

**Authors:** Benjamin Glemain, Charles Assaad, Walid Ghosn, Paul Moulaire, Xavier de Lamballerie, Marie Zins, Gianluca Severi, Mathilde Touvier, Jean-François Deleuze, SAPRIS-SERO study group, Nathanaël Lapidus, Fabrice Carrat

## Abstract

Several studies found an association between the risk of death for COVID-19 patients and hospital overload during the first pandemic wave. We studied this association across the French departments using 82,467 serological samples and a hierarchical Bayesian model. In high-incidence areas, we hypothesized that hospital overload would increase infection fatality rate (IFR) without increasing infection hospitalization rate (IHR). We found that increasing departmental incidence from 3% to 9% rose IFR from 0.42% to 1.14%, and IHR from 1.66% to 3.61%. An increase in incidence from 6% to 12% in people under 60 was associated with an increase in the proportion of people over 60 among those infected, from 11.6% to 17.4%. Higher incidence did increase the risk of death for infected persons, probably due to an older infected population in high-incidence areas rather than hospital overload.

## 1 Introduction

The first wave of the COVID-19 pandemic led to episodes of hospital overload in many parts of the world, requiring the urgent provision of additional hospital beds to prevent excess mortality [1, 2]. Temporary units were set up, but suspected of being less effective than permanent units (although these comparisons were impeded by confounders such as admission criteria) [3–5]. Indeed, hospital overload can manifest in several ways. First, it can appear as a lack of hospital bed availability, resulting in a lower proportion of infected individuals being hospitalized. Second, it can lead to a decrease in the quality of care due to the urgent addition of extra beds with potentially inadequate facilities, equipment, or staff. Hospital overload thus has multiple aspects, making it difficult to measure using a single indicator.

So far, the consequences of these episodes of hospital overload on the risk of death for infected persons have mainly been studied through the number of confirmed COVID-19 cases, using the case fatality rate (CFR) [6–8]. Hospital overload was notably expressed as the ratio between the number of COVID-19 cases and baseline hospital resources, a measure that does not distinguish between situations of high incidence and those of insufficient hospital resources at baseline [6]. In addition, CFR analysis is limited by spatial and temporal variations in the case detection rate (the proportion of new cases that are detected) [9, 10].

To overcome the limitations of the cases-based approach, several studies used serological data and estimated infection fatality rates (IFRs) [11–14]. IFR is defined as the number of deaths attributed to COVID-19 during a given period divided by the number of infected individuals (incidence) over the same period, and corresponds to the risk of death for infected persons. These studies compared IFRs across countries and age groups, but did not explore the impact of hospital overload.

Built on the seroprevalence study SAPRIS-SERO, the present work aimed to estimate the effect of COVID-19 incidence on IFR at the scale of the metropolitan departments of France during the first pandemic wave in the population over 20. We used a Bayesian statistical framework to leverage multiple sources of data and to account for uncertainty surrounding the latent variables when used in regressions (such as incidence and IFR). As incidence (understood as a cumulative incidence over the first wave) was reported to range from 3% to 9% in the French regions (administrative subdivisions gathering several departments), the consequences of an incidence shift from 3% to 9% on IFR were the main object of this study [15, 16].

The relation between incidence and IFR is however confounded, notably because the determinants of IFR may share socio-economic causes with incidence at the scale of departments. Typically, wealthier departments could have a population which travels more (possibly increasing incidence) and which is healthier (decreasing IFR), participating in a spurious negative association between incidence and IFR. Thus, IFR was adjusted (by conditioning and averaging, as described in the Methods section) for the main determinants of COVID-19 outcome: prevalence of diabetes (as a surrogate for obesity), proportion of the population over 60, and number of intensive care beds per inhabitant (see the dedicated subsection: Choice of the covariates) [6, 7, 17, 18]. In the Methods section (equation 15), we formalize the equivalence between (i) the association between incidence and adjusted IFR and (ii) the causal effect of incidence on IFR, using the structural causal models framework.

Finally, the mechanisms by which incidence could influence IFR were explored. Evidence for hospital overload with shortage of hospital beds was sought by examining the effect of COVID-19 incidence on infection hospitalization rate (IHR), which is the proportion of infected individuals being hospitalized for COVID-19. The age of the infected individuals may also play a role in the effect of incidence on IFR. Indeed, it has been suggested that older persons could be under-represented in the population of infected people at the early stages of the epidemic [19]. To determine if infected individuals were actually younger in departments with low incidence, the dependence between COVID-19 incidence in people under 60 and the proportion of individuals over 60 among those infected was investigated.

## 2 Results

### 2.1 Participants

The study included 82,467 persons with a serological test, living in metropolitan France, and over 20 years old. All serological samples were collected between May and November 2020. Among the participants, 319 reported a positive RT-PCR. These latter had a mean age of 52 years, 29% of them were males, and the median time elapsed between RT-PCR and dried blood sampling was 111 days (IQR: 68-128). The participants with a positive RT-PCR were considered infected. On the other hand, 82,148 participants reported no positive RT-PCR (no RT-PCR or a negative RT-PCR). These participants had a mean age of 58 years, and 35% of them were males. A detailed flowchart and a table with the number of samples for each department and age group are provided in the Supplementary information file.

### 2.2 Departmental incidence, IFR, and IHR

The model estimated an incidence of 6.8% over the first wave in metropolitan France, with a 95% credible interval (95% CI) of 6.4 to 7.2%. Figure 1 displays a map of departmental incidence, showing that north-east France was the most affected area. Indeed, departmental incidence ranged from 2.5% (95% CI: 1.8-3.4%), in Ariège (a south-west department), to 13.6% (95% CI: 12.1-15.2%), in Haut-Rhin (a north-east department).

**Fig. 1:**
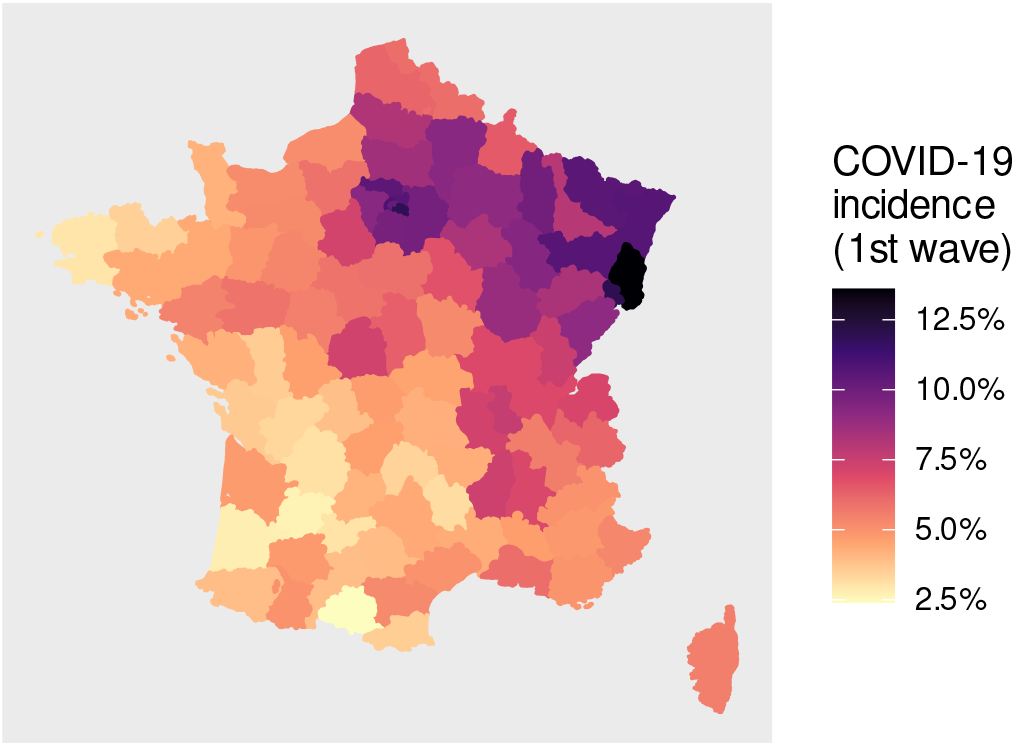
COVID-19 departmental incidence (cumulated over the first wave) in metropolitan France.

The overall infection fatality rate in metropolitan France was 0.94% (95% CI: 0.89-0.99%) and infection hospitalization rate was 3.28% (95% CI: 3.10-3.46%). IFR ranged from 0.27% (95% CI: 0.21-0.33%), in Haute-Garonne (a south-west department with low COVID-19 incidence), to 1.97% (95% CI: 1.74-2.22%), in Haut-Rhin (the department with the highest COVID-19 incidence, located in the north-east of france). IHR ranged from 1.02% (95% CI: 0.72-1.42%), in Tarn-et-Garonne, to 5.92% (95% CI: 5.09-6.79%), in Territoire de Belfort. The Supplementary information file provides exhaustive departmental estimates (incidence, IFR, IHR).

### 2.3 Effect of incidence on IFR and IHR

Figure 2 illustrates the association between departmental incidence and adjusted IFR, or adjusted IHR (adjustment for the proportion of persons over 60 in the population, for the prevalence of diabetes, and for the number of intensive care beds per inhabitant). An incidence of 3% was associated with an adjusted IFR of 0.42% (95% CI: 0.33-0.52%), and an incidence of 9% was associated with an adjusted IFR of 1.14% (95% CI: 0.95-1.39%). The absolute difference (equivalent to the average causal effect of an incidence shift from 3% to 9%) was 0.72% (95% CI: 0.49-1.01%).

**Fig. 2:**
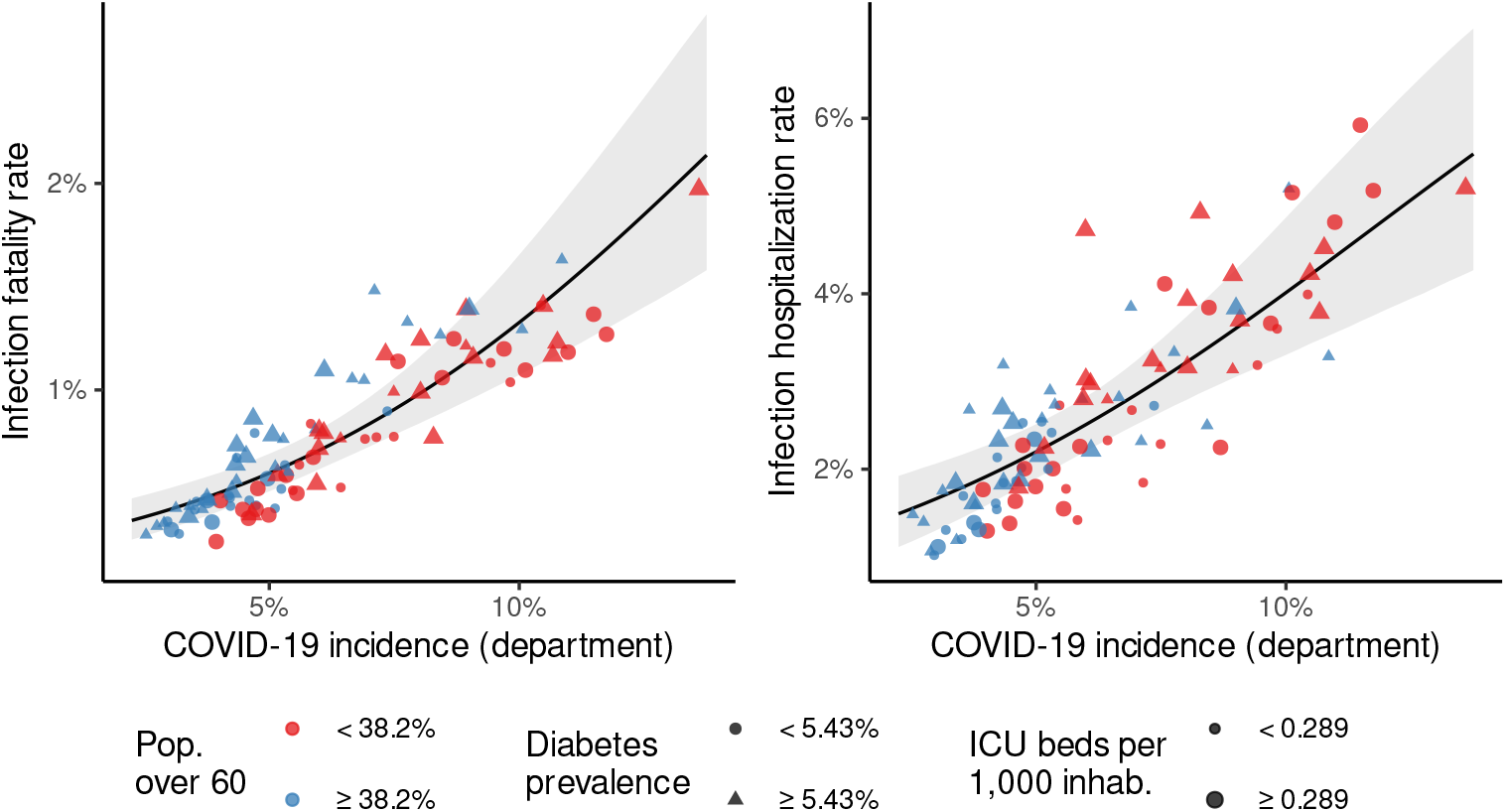
Effect of departmental incidence on infection fatality rate (IFR) and on infection hospitalization rate (IHR). The points represent the mean posterior departmental estimates. Black line and gray zone: Posterior mean and 95% CI of the expected adjusted departmental IFR given incidence (or expected causal effect of incidence on IFR: see equation 15 of the Methods). The same description applies to IHR. The covariates are represented relative to their medians. Pop. over 60: Proportion of adult population over 60. ICU beds per 1,000 inhab.: Number of intensive care beds per 1,000 inhabitants.

An incidence of 3% was associated with an adjusted IHR of 1.66% (95% CI: 1.30-2.06%), and an incidence of 9% was associated with an adjusted IHR of 3.61% (95% CI: 3.05-4.28%). The absolute difference was 1.94% (95% CI: 1.18-2.80%).

Complementary results (univariate analysis, role of the confounders) are provided in the Supplementary information file.

### 2.4 Association between incidence and age of infected persons

As illustrated in Figure 3, a shift in incidence in the persons under 60 from 6% to 12% (typical observed values) was associated with an increase in the expected proportion of persons over 60 among those infected for a department with the same age structure as metropolitan France, from 11.6% (95% CI: 9.6-13.6%) to 17.4% (95% CI: 15.5-19.5%). The absolute difference was 5.8% (95% CI: 2.9-8.8%).

**Fig. 3:**
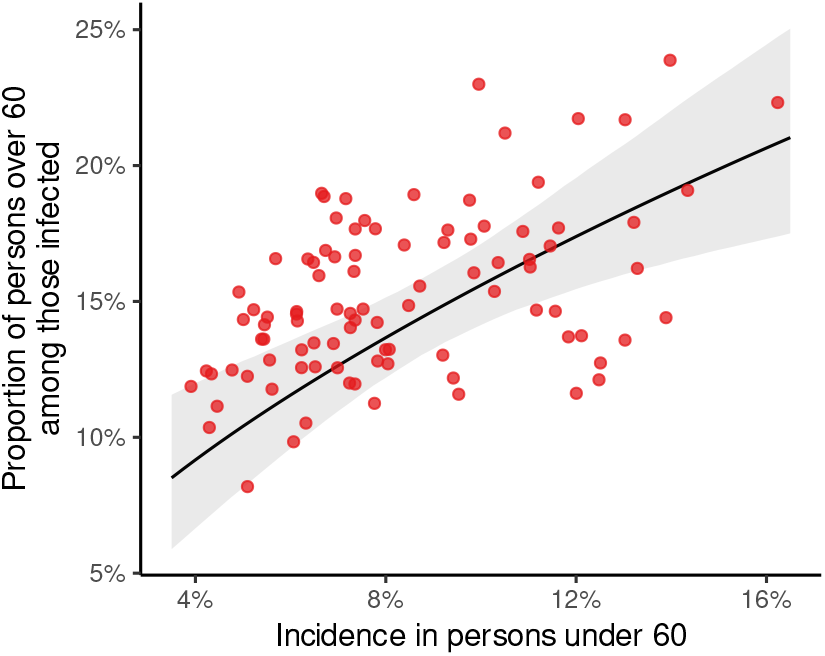
Association between incidence in people under 60 and the proportion of people over 60 among those infected. The points represent the mean posterior departmental estimates. Black line and gray zone: Posterior mean and 95% CI of the expected proportion of persons over 60 among those infected (for a department with the same age structure as metropolitan France).

## 3 Discussion

This study explored the role of hospital overload on the risk of death for COVID-19 patients (IFR) using data collected in France following the first pandemic wave. We found that a higher departmental incidence was associated with a higher adjusted IFR, corresponding to a causal effect of incidence on IFR. This effect could possibly be explained by the age of the infected persons, as we found a higher proportion of people aged over 60 among those infected in high-incidence departments. The role of hospital overload was explored through the analysis of the probability of hospitalization when infected (IHR). In case of hospital overload with a lack of beds, we would have expected a decrease in the proportion of infected individuals who are hospitalized. In case of hospital overload with sufficient additional beds available but a decrease in the quality of care, we would have expected IHR to remain stable with incidence (since infected individuals could have been hospitalized without restriction). On the contrary, we found that an incidence shift from 3% to 9% increased IHR by the same magnitude as IFR (by a factor of two to three), consistent with the increase in the age of infected individuals with incidence (since both IHR and IFR increase with age) [20]. Previous studies have compared IFR between countries and have found that age-specific IFR was the main predictor of country-level IFR [11]. However, IFR varied strongly between countries (by a factor of more than 30 in [11]), even after accounting for the age of the infected individuals [13, 14]. In particular, European countries faced higher IFRs than expected [11–13], suggesting the importance of other determinants of IFR than age and health-care capacity [11]. Focusing on a single country was a first strength of our study, as it eliminated the influence of any country-level confounder (heterogeneity in populations, healthcare systems, mitigation policies, etc.), by design. Furthermore, the association between departmental incidence and IFR was not explained by the covariates we used: baseline hospital resources (measured as the number of intensive care beds per inhabitant), proportion of the population over 60, or prevalence of diabetes (used as a proxy for obesity). This association between incidence and adjusted IFR was also suggested in one international comparison (see the Figure 3 of [14]).

Some other studies found a positive association between a higher case fatality rate (CFR) and hospital overload in the US and in France [4, 6]. In these studies, hospital overload was measured either by the number of hospitalizations for COVID-19 or by the ratio between COVID-19 cases and hospital resources, such as ICU beds or nursing staff. In one of these studies, CFR was standardized for the age of the population, but not for the age of the infected individuals [4]. These findings may be explained by the role of incidence. Indeed, hospital overload increases with incidence, as does the age of infected individuals (and consequently, IFR). Incidence could therefore be a confounder for hospital overload and IFR.

An important strength of this study was the Bayesian statistical framework, which accounted for uncertainty in the latent variables used in the regressions (such as incidence or IFR). Indeed, underestimation of incidence results in overestimation of IFR (since the number of COVID-19 related deaths is known), and overestimation of incidence leads to underestimation of IFR. Thus, sampling variation contributes to a spurious negative association between incidence and IFR, as evidenced in the posterior predictive checks (see the Supplementary information file). Considering these latent variables as known during the regression step would therefore have led to biased results.

A limitation of our study was that focusing on a single country could reduce external validity, particularly for countries on other continents or with different levels of development. A second limitation relates to the causal assumptions we have made, as they result in a simplified view of the relation between the variables. We used the prevalence of diabetes as a surrogate for obesity because it is easier to collect data on (through diabetes medication), and not all risk factors for COVID-19 death were considered (like immunosuppression). However, these assumptions enabled formal causal reasoning using graphs, which is not yet widely practiced in health research [21]. This formalism contributes to greater clarity regarding the questions asked and the mechanisms under study.

In conclusion, this study found that a higher incidence increased the risk of death among individuals infected with COVID-19. However, the mechanism of this effect could be related to a shift in the profile of infected individuals with incidence. Indeed, the departments with lower incidence tended to have a younger infected population (to be distinguished from the age of the general population of the department). These findings prompt a reinterpretation of studies that have observed an association between the number of COVID-19 cases in a specific location and the risk of death among infected individuals, as the age of infected persons was generally not considered. It is important to emphasize that while we did not find epidemiological evidence implicating hospital overload in increasing the risk of death among COVID-19 patients, this does not negate the possibility of such an effect.

## 4 Methods

### 4.1 SAPRIS-SERO study

This study used the data of the SAPRIS-SERO serosurvey, previously described [16, 22, 23]. SAPRIS-SERO was built on SAPRIS (“SAnté, Perception, pratiques, Relations et Inégalités Sociales en population générale pendant la crise COVID-19”), a cohort whose inclusions began in March 2020, which studied epidemiological and sociological aspects of the COVID-19 epidemic in France [22]. The adult participants of SAPRIS were recruited from three cohorts based on the general population (without particular selection on a disease):

- The cohort NutriNet-Santé focused on nutrition, with online follow-up. It included 170,000 participants at the start of the study in 2009 [24].
- The cohort CONSTANCES was set up in 2012 and included 204,973 adults, selected to be a representative sample of the French adult population [25].
- E3N/E4N is a multi-generational adult cohort including 113,000 persons: the women recruited at the start of the study (1990), their children, and the fathers of these children [26].

All participants from the initial cohorts who had regular internet access and were still being followed in 2020 were invited to participate in the SAPRIS study, which involved self-administered questionnaires during the first wave. These questionnaires covered demographic information and the history of SARS-CoV-2 testing by RT-PCR. A total of 93,610 SAPRIS participants were over 20, completed the questionnaires, and resided in metropolitan France. These participants were then invited to join the SAPRISSERO study by collecting a single dried-blood spot sample themselves. The samples were sent to a virology laboratory (Unité des virus émergents, Marseille, France) for serological analysis using the commercial ELISA test (Euroimmun, Lübeck, Germany), which detects anti-SARS-CoV-2 IgG antibodies targeting the S1 domain of the spike protein. The ELISA assays performed on dried-blood spot samples demonstrated a sensitivity of 98.1% to 100% and a specificity of 99.3% to 100% when compared to conventional serum assays as a standard [27, 28].

### 4.2 External data

The results of two independent French seroprevalence studies were used as prior distributions for national seroprevalence (metropolitan France) [15, 29]. The results of a diagnostic study concerning the ELISA test (IgG anti-S1 from Euroimmun) were used as prior distributions for sensitivity and specificity [30].

The French population structure by age and by administrative department came from the census of January first, 2020 (Insee, Institut national de la statistique et des études économiques) [31]. The data about COVID-19-related hospitalizations during the first semester (before July first, 2020) by administrative department were obtained from the SI-VIC database, the exhaustive national inpatient surveillance system used during the pandemic [32]. The data about general population mortality (including deaths occurring in nursing homes) attributed to COVID-19 during the first semester (before July first, 2020) were obtained from the CépiDc (Centre d’épidémiologie sur les causes médicales de décès), online (open data) or directly [33]. Raw diabetes prevalence in French departments in 2019 (pre-pandemic) was provided by Santé Publique France (open data), based on an exhaustive monitoring of anti-diabetic drugs use (Système national des données de santé) [34]. The number of ICU (intensive care unit) beds per inhabitant in 2019 (pre-pandemic) was obtained from the DREES (Direction de la recherche, des études, de l’évaluation et des statistiques) [35].

### 4.3 Choice of the covariates

Figure 4 features a causal graph representing departmental incidence (*X*), IFR or IHR (*Y*), unobserved socio-economic variables (*C*), and the determinants of COVID-19 outcome (according to [6, 7, 17, 18]). Our most critical assumptions, which are included in the graph, are:

**Fig. 4:**
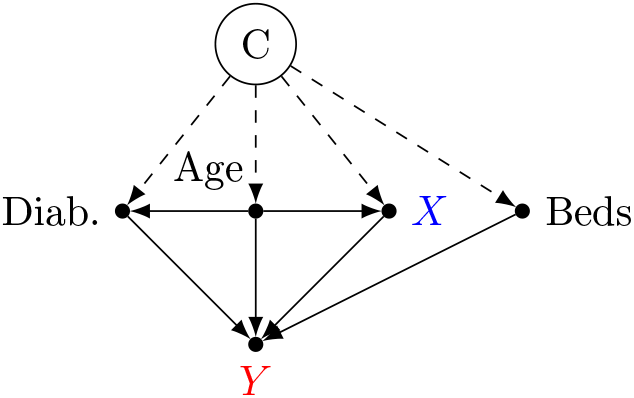
Causal graph. The variables are considered at the departmental scale. The effect of *X* on *Y* can be estimated by adjusting on {Diab., Age, Beds}. *X*: COVID-19 incidence. *Y* : IFR (infection fatality rate) or IHR (infection hospitalization rate). Diab.: Prevalence of diabetes. Beds: Number of intensive care beds per inhabitant. Age: Proportion of population over 60. Dashed arrows represent the effects of unmeasured confounders C.

- Age and diabetes are the main individual risk factors for COVID-19 severity, influencing hospitalization and mortality.
- The determinants of IFR and IHR at the departmental scale act through the prevalence of these individual risk factors, through incidence, or through the number of intensive care beds per inhabitant (the latter having a potentially decisive role for IFR but acting as a surrogate for hospital beds when considering IHR).

Given this causal graph (Figure 4), the set {Diab., Age, Beds} satisfies the back-door criterion relative to (*X, Y*) and allows for estimating the causal effect of *X* on *Y* despite the presence of unobserved socio-economic variables, using the back-door adjustment formula (equation 15) [36]. Prevalence of diabetes was chosen as a surrogate for obesity because it is easier to quantify precisely (through data on the sale of diabetes medication).

### 4.4 Statistical model

The statistical analysis was carried out within a Bayesian framework. An overview of the model is featured in Figure 5, where the equations of the model are referenced next to the variables. In the remainder of this section, prior distributions are not always explicitly written. If so, the latter are uniform. Age groups are indexed by the letter *i* (*i* = 0 for individuals aged 20 to 59, and *i* = 1 for individuals aged over 60). The departments are indexed by the letter *j*, ranging from 1 to 95.

**Fig. 5:**
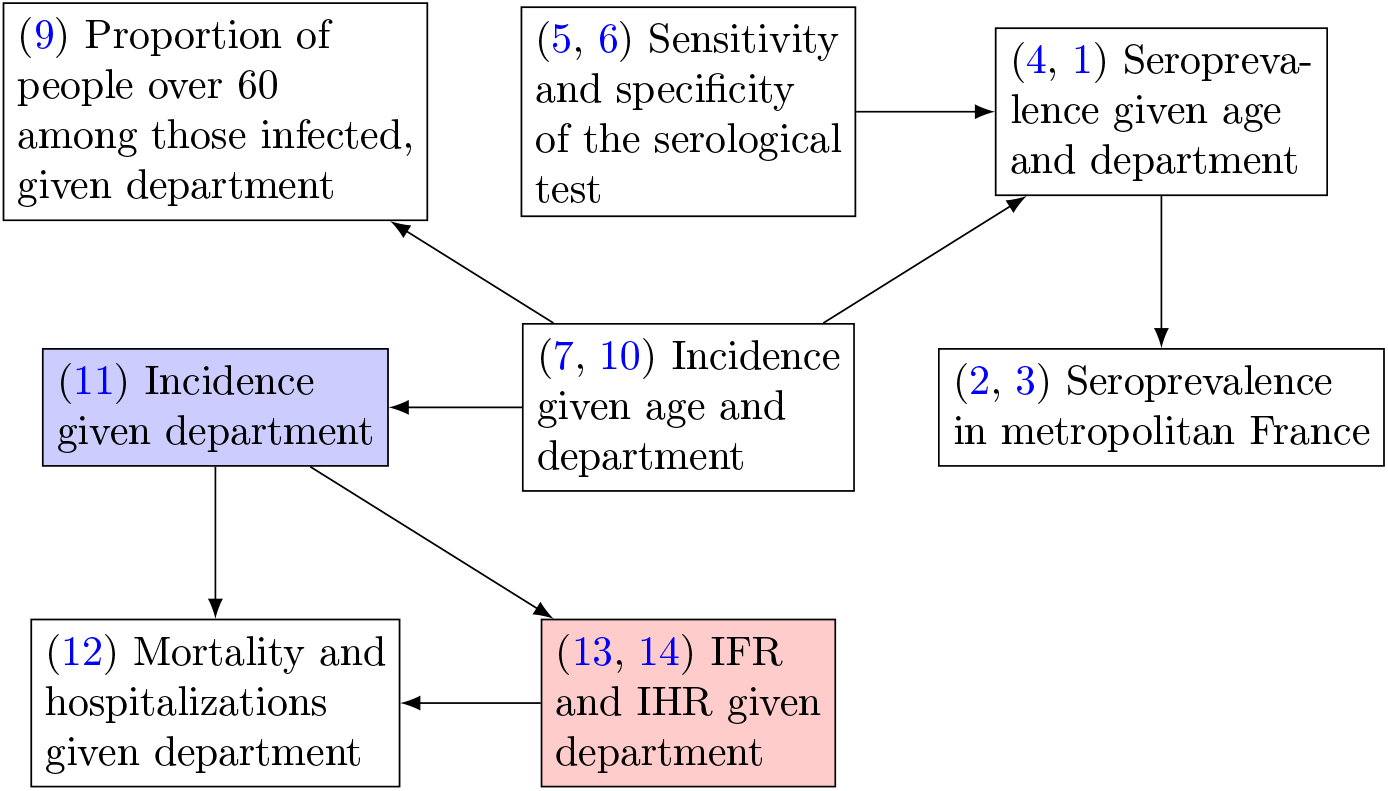
Overview of the model. The blue and red rectangles represent the exposure and outcome of the main analysis, respectively. The numbers indicate the equations associated with the variables (see the Model section). IFR: Infection fatality rate. IHR: Infection hospitalization rate.

For an age group *i* and a department *j*, the participants without a positive RT-PCR nor missing data on department contributed to the estimation of seroprevalence *s*_*i,j*_, considering *N*_*i,j*_ the number of these participants, and *y*_*i,j*_ the number of positive serological tests:

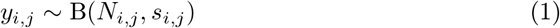

Seroprevalence at the scale of metropolitan France (sero_France_) was obtained by post-stratification from *s*_*i,j*_ and pop_*i,j*_, the size of the population corresponding to this group:

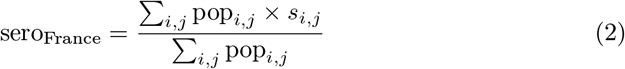

Seroprevalence estimates from other surveys were incorporated using beta distributions:

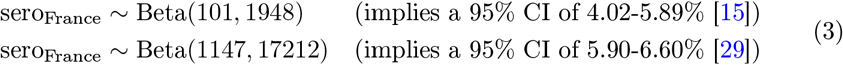

For an age group *i* and a department *j*, incidence (cumulated over the first semester) was denoted *p*_*i,j*_. Seroprevalence *s*_*i,j*_ was linked to incidence *p*_*i,j*_ and to the sensitivity (Se) and specificity (Sp) of the serological test:

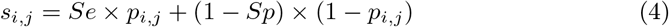

Prior distributions for sensitivity and specificity originated from [30]:

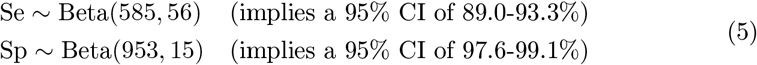

The participants with a positive RT-PCR contributed to the likelihood of sensitivity. With *N*_se_ and *y*_se_ the number of total and positive (respectively) serological tests in this group,

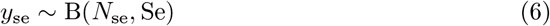

Incidence *p*_*i,j*_ was modeled on the logit scale. As it was suggested that older persons could be under-represented in the population of infected persons at the early stages of the epidemic [19], every department had a unique log-odds ratio for age over 60 (*β*_*j*_), possibly influenced by its intercept *α*_*j*_ (logit of incidence in the 20-59 in the department *j*) through a linear regression with intercept *μ*_age_, slope *b*_age_ and standard deviation *σ*_age_ (hierarchical modeling, *β*_*j*_ is a random effect):

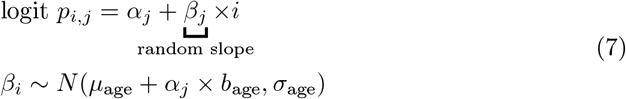

The *α*_*j*_ departmental intercepts entailed spatial auto-correlation through an ICAR (intrinsic conditional auto-regressive) component *ϕ*_*j*_ (as described and implemented in this reference [37]), associated with an overall intercept *μ*_*α*_ and a scale parameter *σ*_*ϕ*_ representing the amount of spatial correlation:

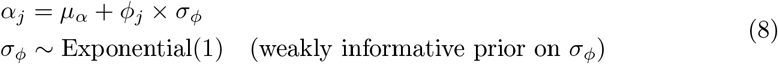

The proportion age_infected,j_ of persons over 60 among those infected in a department *j* was reconstructed from *p*_*i,j*_ and from 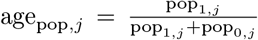 (the proportion of persons above 60 in the population of the department *j*):

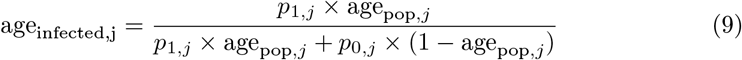

For a given incidence in the persons under 60, we estimated an expected proportion of persons over 60 among those infected for a department with the same age structure as metropolitan France. This expected proportion was reconstructed from the coefficients *μ*_*age*_, *β*_*age*_, and *σ*_*age*_ (equation 7), according to a procedure described in the Supplementary information file (“Computation of expectations” section). This analysis aimed to illustrate the dependence between incidence in people under 60 and incidence in those over 60, and its possible consequences for IFR.

The *K* participants with a positive RT-PCR and no missing data concerning the department contributed directly to incidence. With IS_*k*_ being the infection status of the participant *k* (IS_*k*_ is always equal to 1 in this positive RT-PCR group),

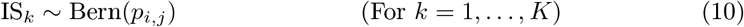

Departmental incidence inc_*j*_ was obtained by post-stratification from *p*_*i,j*_ and pop_*i,j*_:

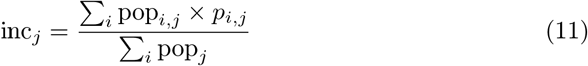

For a department *j*, the counts of deaths (D_*j*_) and hospitalizations (H_*j*_) of the first semester were modeled with Poisson regressions:

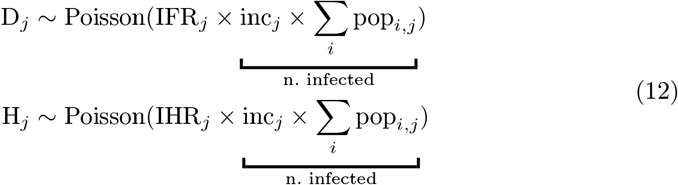

Departmental IFRs and IHRs were modeled as logistic functions of linear predictors *g*_IFR_(*j*) and *g*_IHR_(*j*), ranging respectively from 0% to 5% and from 0% to 10%:

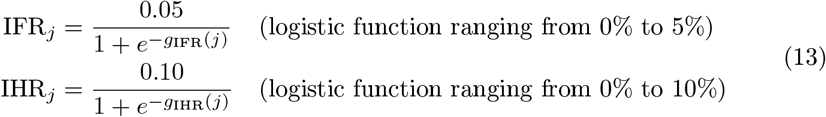

These linear predictors included a departmental random intercept, a slope representing the role of incidence, and coefficients associated with the covariates (prevalence of diabetes, number of intensive care beds per inhabitant and proportion of the population over 60). With diab_*j*_, age_pop,*j*_, and beds_*j*_ being the covariates for the department *j*,

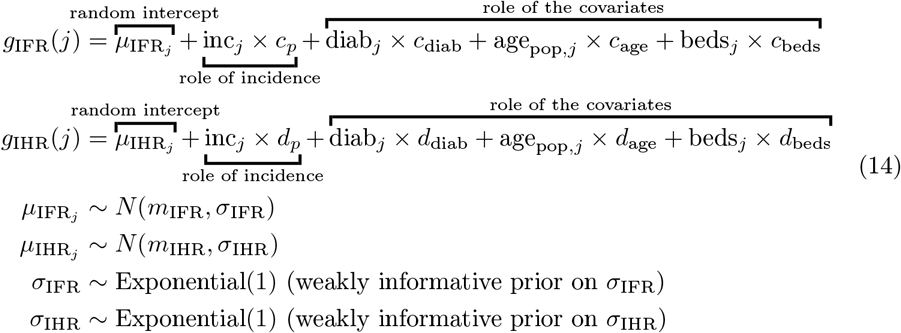

Because the covariates satisfy the back-door criterion, the adjusted IFR can be equated to the causal effect of incidence on IFR using the back-door adjustment formula [36]. *P* (IFR|*do*(inc = *x*)) denotes this causal effect, which is the distribution of departmental IFRs if incidence was artificially set to the value *x* without any other modification. With *z*_*j*_ denoting the vector of covariates for department *j*, and because the distribution of these covariates in the French departments is known 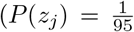 for all *j*),

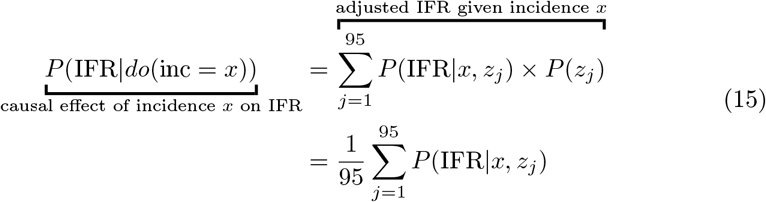

The coefficients of equation 14 were used to compute *E*[IFR|*do*(inc = *x*)], as described in the supplementary information file (“Reconstruction of expectations” section). The target of this study was the average causal effect of an incidence shift from 3% to 9%:

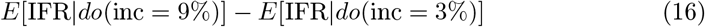

This average causal effect corresponds to the expected difference in IFR when artificially setting incidence to 9% versus 3% in a department of metropolitan France (without changing anything else than incidence). The same procedure was applied to estimate the average causal effect of this incidence shift on IHR.

### 4.5 Algorithm and software

The data management was done using R version 4.3.1, and the modeling was performed with Stan (R package cmdstanr version 0.5.3), which implements Hamiltonian Monte Carlo (HMC) [38, 39]. The models for the random coefficients (*β*_*j*_, 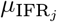 and 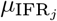) employed non-centered parameterizations to improve HMC convergence [40]. The Monte Carlo sampling consisted of 8 chains of 2,000 iterations each (including 1,000 warm up iterations). Trace plots, 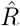 statistics and effective Monte Carlo sample sizes provided by Stan were used to assess convergence. Posterior predictive checks are provided in the Supplementary information file. The model’s code (in Stan) is provided in the Supplementary information file and in a GitHub repository (https://github.com/bglemain/does-hospital-overload).

### 4.6 Ethical approval and consent to participate

Ethical approval and written or electronic informed consent were obtained from each participant before enrollment in the original cohort. The SAPRIS-SERO study was approved by the Sud-Mediterranée III ethics committee (approval 20.04.22.74247) and electronic informed consent was obtained from all participants for dried blood spot testing. The study was registered (#NCT04392388). All methods were performed in accordance with the relevant guidelines and regulations.

## Data Availability

In regard to data availability, data from the study are protected under the protection of health data regulation set by the French National Commission on Informatics and Liberty (Commission Nationale de l Informatique et des Libertes, CNIL). The data can be made available upon reasonable request to F.C., after a consultation with the steering committee of the SAPRIS-SERO study. The French law forbids us to provide free access to SAPRIS-SERO data; access could however be given by the steering committee after legal verification of the use of the data. Please, feel free to come back to us should you have any additional question.

## Supplementary information

The Supplementary information file attached with this article contains:

- A flowchart
- The number of serological samples for each department and age group
- The exhaustive list of departmental estimates (incidence, IFR, IHR) and characteristics (covariates, population size, counts of deaths and hospitalizations)
- Complementary results (univariate analysis, role of the confounders)
- The procedure used to compute expectations after a logistic transformation
- Posterior predictive checks
- The Stan code

## Acknowledgments

The authors warmly thank all the volunteers of the Constances, E3N–E4N, and NutriNet-Santé cohorts. We thank the staff of the Constances, E3N–E4N and NutriNet-Santé cohorts that have worked with dedication and engagement to collect and manage the data used for this study and to ensure continuing communication with the cohort participants. We thank the CEPH-Biobank staff for their adaptability and the quality of their work. We thank all the members of the SAPRIS-SERO study group (see Supplementary information file).

## Declarations

### Ethics approval

Ethical approval and written or electronic informed consent were obtained from each participant before enrolment in the original cohort. The SAPRIS-SERO study was approved by the Sud-Mediterranée III ethics committee (approval #20.04.22.74247)

### Consent to participate

An electronic informed consent was obtained from all participants for DBS testing

### Consent for publication

Participants can not be identified on the basis of this article

### Availability of data and materials

In regard to data availability, data from the study are protected under the protection of health data regulation set by the French National Commission on Informatics and Liberty (Commission Nationale de l’Informatique et des Libertés, CNIL). The data can be made available upon reasonable request to F.C., after a consultation with the steering committee of the SAPRIS-SERO study. The French law forbids us to provide free access to SAPRIS-SERO data; access could however be given by the steering committee after legal verification of the use of the data. Please, feel free to come back to us should you have any additional question.

### Code availability

The code Stan for the model is provided in Supplementary information file

### Authors’ contributions

F.C., N.L., C.A., W.G., P.M, and B.G. conceived and designed the study. B.G. implemented the model and wrote the manuscript. All the authors reviewed and edited the manuscript.

## Consortium (SAPRIS-SERO study group)

Fabrice Carrat^1,2^, Pierre-Yves Ancel^11^, Marie-Aline Charles^11^, Gianluca Severi^7,8^, Mathilde Touvier^9^, Marie Zins^5,6^, Sofiane Kab^6^, Adeline Renuy^6^, Stephane Le-Got^6^, Celine Ribet^6^, Mireille Pellicer^6^, Emmanuel Wiernik^6^, Marcel Goldberg^6^, Fanny Artaud^7^, Pascale Gerbouin-Rérolle^7^, Mélody Enguix^7^, Camille Laplanche^7^, Roselyn Gomes-Rima^7^, Lyan Hoang^7^, Emmanuelle Correia^7^, Alpha Amadou Barry^7^, Nadège Senina^7^, Julien Allegre^9^, Fabien Szabo de Edelenyi^9^, Nathalie Druesne-Pecollo^9^, Younes Esseddik^9^, Serge Hercberg^9^, Mélanie Deschasaux^9^, Marie-Aline Charles^11^, Valérie Benhammou^12^, Anass Ritmi^13^, Laetitia Marchand^13^, Cecile Zaros^13^, Elodie Lordmi^13^, Adriana Candea^13^, Sophie de Visme^13^, Thierry Simeon^13^, Xavier Thierry^13^, Bertrand Geay^13^, Marie-Noelle Dufourg^13^, Karen Milcent^13^, Delphine Rahib^14^, Nathalie Lydie^14^, Clovis Lusivika-Nzinga^1^, Gregory Pannetier^1^, Nathanael Lapidus^1,2^, Isabelle Goderel^1^, Céline Dorival^1^, Jérôme Nicol^1^, Olivier Robineau^1^, Cindy Lai^15^, Liza Belhadji^15^, Hélène Esperou^15^, Sandrine Couffin-Cadiergues^15^, Jean-Marie Gagliolo^16^, Hélène Blanché^10^, Jean-Marc Sébaoun^10^, Jean-Christophe Beaudoin^10^, Laetitia Gressin^10^, Valérie Morel^10^, Ouissam Ouili^10^, Jean-François Deleuze^10^, Laetitia Ninove^4^, Stéphane Priet^4^, Paola Mariela Saba Villarroel^4^, Toscane Fourié^4^, Souand Mohamed Ali^4^, Abdenour Amroun^4^, Morgan Seston^4^, Nazli Ayhan^4^, Boris Pastorino^4^, Xavier de Lamballerie^4^.

^1^Sorbonne Université, Inserm, Institut Pierre-Louis d’épidémiologie et de santé publique, Paris, France

^2^Département de santé publique, Hôpital Saint-Antoine, AP-HP.Sorbonne université, Paris, France

^4^Unité des Virus Émergents, UVE, Aix Marseille Univ, IRD 190, INSERM 1207, IHU Méditerranée Infection, Marseille, France

^5^Paris University, Paris, France

^6^Université Paris-Saclay, Université de Paris, UVSQ, Inserm UMS 11, Villejuif, France

^7^CESP UMR1018, Université Paris-Saclay, UVSQ, Inserm, Gustave Roussy, Villejuif, France

^8^Department of Statistics, Computer Science and Applications, University of Florence, Italy

^9^Sorbonne Paris Nord University, Inserm U1153, Inrae U1125, Cnam, Nutritional Epidemiology Research Team (EREN), Epidemiology and Statistics Research Center – University of Paris (CRESS), Bobigny, France

^10^Fondation Jean Dausset-CEPH (Centre d’Etude du Polymorphisme Humain), CEPH-Biobank, Paris, France

^11^Centre for Research in Epidemiology and StatisticS (CRESS), Inserm, INRAE, Université de Paris, Paris, France

^12^EPIPAGE-2 Joint Unit, Paris, France

^13^ELFE Joint Unit, Paris, France

^14^Santé Publique France, Paris, France

^15^Inserm, Paris, France

^16^Aviesan, Inserm, Paris, France

## References

[1] Candel, F. J. et al. Temporary hospitals in times of the COVID pandemic. An example and a practical view. Rev. Esp. Quimioter. 34, 280 (2021).

[2] Lefrant, J.-Y. et al. A national healthcare response to intensive care bed requirements during the COVID-19 outbreak in France. Anaesth Crit Care Pain Med 39, 709–715 (2020).

[3] Jimenez, J. V. et al. Outcomes in Temporary ICUs Versus Conventional ICUs: An Observational Cohort of Mechanically Ventilated Patients With COVID-19– Induced Acute Respiratory Distress Syndrome. Crit Care Explor 4, e0668 (2022).

[4] Souris, M. & Gonzalez, J.-P. COVID-19: Spatial analysis of hospital case-fatality rate in France. PLoS One 15, e0243606 (2020).

[5] Zappella, N. et al. Temporary ICUs during the COVID-19 pandemic first wave: Description of the cohort at a French centre. BMC Anesthesiol 22, 310 (2022).

[6] Janke, A. T. et al. Analysis of Hospital Resource Availability and COVID-19 Mortality Across the United States. J Hosp Med 16, 211–214 (2021).

[7] Sen-Crowe, B., Sutherland, M., McKenney, M. & Elkbuli, A. A Closer Look Into Global Hospital Beds Capacity and Resource Shortages During the COVID-19 Pandemic. J Surg Res 260, 56–63 (2021).

[8] Dudel, C. et al. Monitoring trends and differences in COVID-19 case-fatality rates using decomposition methods: Contributions of age structure and age-specific fatality. PLoS One 15, e0238904 (2020).

[9] Lau, H. et al. Evaluating the massive underreporting and undertesting of COVID-19 cases in multiple global epicenters. Pulmonology 27, 110–115 (2021).

[10] Borgdorff, M. W. New Measurable Indicator for Tuberculosis Case Detection. Emerg. Infect. Dis. 10, 1523 (2004).

[11] COVID-19 Forecasting Team. Variation in the COVID-19 infection-fatality ratio by age, time, and geography during the pre-vaccine era: A systematic analysis. Lancet 399, 1469–1488 (2022).

[12] O’Driscoll, M. et al. Age-specific mortality and immunity patterns of SARS-CoV-2. Nature 590, 140–145 (2021).

[13] Pezzullo, A. M., Axfors, C., Contopoulos-Ioannidis, D. G., Apostolatos, A. & Ioannidis, J. P. A. Age-stratified infection fatality rate of COVID-19 in the non-elderly population. Environ Res 216, 114655 (2023).

[14] Ioannidis, J. P. A. Infection fatality rate of COVID-19 inferred from seroprevalence data. Bull World Health Organ 99, 19–33F (2021).

[15] Le Vu, S. et al. Prevalence of SARS-CoV-2 antibodies in France: Results from nationwide serological surveillance. Nat Commun 12, 3025 (2021).

[16] Carrat, F. et al. Antibody status and cumulative incidence of SARS-CoV-2 infection among adults in three regions of France following the first lockdown and associated risk factors: A multicohort study. Int J Epidemiol 50, 1458–1472 (2021).

[17] COVID-ICU Group on behalf of the REVA Network and the COVID-ICU Investigators. Clinical characteristics and day-90 outcomes of 4244 critically ill adults with COVID-19: A prospective cohort study. Intensive Care Med 47, 60–73 (2021).

[18] Brown, P. A. Country-level predictors of COVID-19 mortality. Sci Rep 13, 9263 (2023).

[19] Tran Kiem, C. et al. SARS-CoV-2 transmission across age groups in France and implications for control. Nat Commun 12, 6895 (2021).

[20] Glemain, B. et al. Estimating SARS-CoV-2 infection probabilities with serological data and a Bayesian mixture model. Sci Rep 14, 9503 (2024).

[21] Tennant, P. W. G. et al. Use of directed acyclic graphs (DAGs) to identify confounders in applied health research: Review and recommendations. Int J Epidemiol 50, 620–632 (2021).

[22] Carrat, F. et al. Incidence and risk factors of COVID-19-like symptoms in the French general population during the lockdown period: A multi-cohort study. BMC Infect Dis 21, 169 (2021).

[23] Carrat, F. et al. Age, COVID-19-like symptoms and SARS-CoV-2 seropositivity profiles after the first wave of the pandemic in France. Infection 50, 257–262 (2022).

[24] Hercberg, S. et al. The Nutrinet-Santé Study: A web-based prospective study on the relationship between nutrition and health and determinants of dietary patterns and nutritional status. BMC Public Health 10, 242 (2010).

[25] Zins, M. & Goldberg, M. The French CONSTANCES population-based cohort: Design, inclusion and follow-up. Eur J Epidemiol 30, 1317–1328 (2015).

[26] Clavel-Chapelon, F. & E3N Study Group. Cohort Profile: The French E3N Cohort Study. Int J Epidemiol 44, 801–809 (2015).

[27] Morley, G. L. et al. Sensitive Detection of SARS-CoV-2-Specific Antibodies in Dried Blood Spot Samples. Emerg Infect Dis 26, 2970–2973 (2020).

[28] Zava, T. T. & Zava, D. T. Validation of dried blood spot sample modifications to two commercially available COVID-19 IgG antibody immunoassays. Bioanalysis 13, 13–28 (2021).

[29] Warszawski, J. et al. Trends in social exposure to SARS-Cov-2 in France. Evidence from the national socio-epidemiological cohort–EPICOV. PLoS One 17, e0267725 (2022).

[30] Otter, A. D. et al. Implementation and Extended Evaluation of the Euroimmun Anti-SARS-CoV-2 IgG Assay and Its Contribution to the United Kingdom’s COVID-19 Public Health Response. Microbiol Spectr 10, e0228921 (2022).

[31] Populations légales 2020 Recensement de la population Régions, départements, arrondissements, cantons et communes. https://www.insee.fr/fr/statistiques/6683031?sommaire=6683037.

[32] Données hospitalières relatives à l’épidémie de COVID-19 (SIVIC). https://www.data.gouv.fr/fr/datasets/donnees-hospitalieres-relatives-a-lepidemie-de-covid-19/.

[33] Covid-19 - Inserm-CépiDc. https://opendata.idf.inserm.fr/cepidc/covid-19/.

[34] Géodes - Santé publique France. https://geodes.santepubliquefrance.fr/#c=home.

[35] La Statistique annuelle des établissements (SAE) | Direction de la recherche, des études, de l’évaluation et des statistiques. https://drees.solidarites-sante.gouv.fr/sources-outils-et-enquetes/00-la-statistique-annuelle-des-etablissements-sae.

[36] Pearl, J. Causal Diagrams for Empirical Research. Biometrika 82, 669–688 (1995).

[37] Morris, M. et al. Bayesian hierarchical spatial models: Implementing the Besag York Mollié model in stan. Spat Spatiotemporal Epidemiol 31, 100301 (2019).

[38] R Core Team. R: A Language and Environment for Statistical Computing. R Foundation for Statistical Computing, Vienna, Austria (2023). URL https://www.R-project.org/.

[39] Carpenter, B. et al. Stan: A Probabilistic Programming Language. J Stat Softw 76, 1 (2017).

[40] Papaspiliopoulos, O. & Roberts, G. Non-Centered Parameterisations for Hierarchical Models and Data Augmentation. Bayesian Statistics 7, 307–326 (2003).

