## Supplementary information for "Does hospital overload increase the risk of death when infected by SARS-CoV-2?"

### Does hospital overload increase the risk of death for COVID-19 patients?

**Posterior predictive checks** **17**

**Stan code** **22**

#### Flowchart

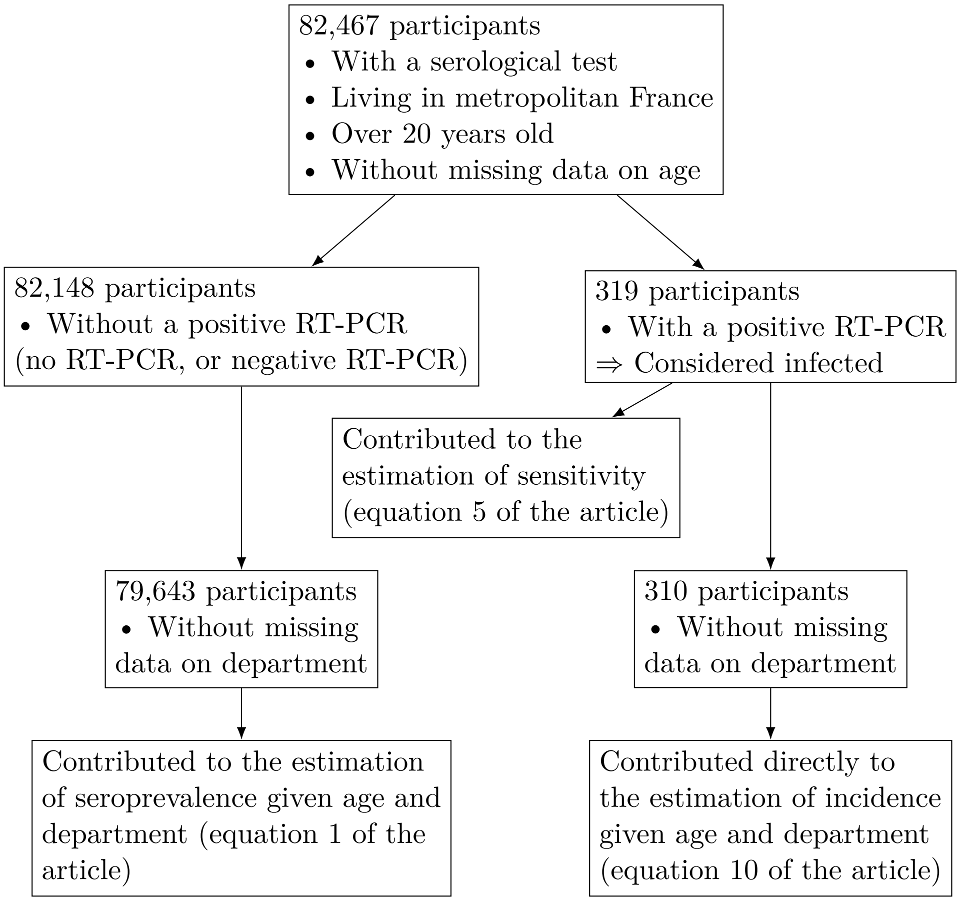

Figure 1: Flowchart of the participants of SAPRIS-SERO

#### Number of serological samples for each department and age group

Table 1: Number of serological samples for each department and age group

| | | Age group | |
|---|---|---|---|
| Code | Depatment | Under 60 | Over 60 |
| 1 | Ain | 244 | 120 |
| 2 | Aisne | 66 | 65 |
| 3 | Allier | 63 | 62 |
| 4 | Alpes De Haute Provence | 43 | 59 |
| 5 | Hautes Alpes | 42 | 46 |
| 6 | Alpes Maritimes | 256 | 240 |
| 7 | Ardeche | 91 | 74 |
| 8 | Ardennes | 41 | 30 |
| 9 | Ariege | 36 | 33 |
| 10 | Aube | 60 | 109 |
| 11 | Aude | 67 | 160 |
| 12 | Aveyron | 64 | 130 |
| 13 | Bouches Du Rhone | 1265 | 1381 |
| 14 | Calvados | 648 | 605 |
| 15 | Cantal | 35 | 49 |
| 16 | Charente | 623 | 752 |
| 17 | Charente Maritime | 156 | 451 |
| 18 | Cher | 68 | 133 |
| 19 | Correze | 62 | 93 |
| 20 | Corse | 38 | 47 |
| 21 | Cote D Or | 213 | 354 |
| 22 | Cotes D Armor | 890 | 954 |
| 23 | Creuse | 13 | 48 |
| 24 | Dordogne | 83 | 208 |
| 25 | Doubs | 182 | 279 |
| 26 | Drome | 167 | 301 |
| 27 | Eure | 91 | 197 |
| 28 | Eure Et Loir | 88 | 138 |
| 29 | Finistere | 302 | 477 |
| 30 | Gard | 633 | 742 |
| 31 | Haute Garonne | 2147 | 1585 |
| 32 | Gers | 57 | 98 |
| 33 | Gironde | 1869 | 1570 |

Table 1: Number of serological samples for each department and age group *(continued)*

| Code | Depatment | Age group<br>Under 60 | Over 60 |
|---|---|---|---|
| 34 | Herault | 386 | 664 |
| 35 | Ille Et Vilaine | 1512 | 988 |
| 36 | Indre | 35 | 78 |
| 37 | Indre Et Loire | 1162 | 1198 |
| 38 | Isere | 639 | 711 |
| 39 | Jura | 69 | 121 |
| 40 | Landes | 104 | 235 |
| 41 | Loir Et Cher | 88 | 155 |
| 42 | Loire | 199 | 321 |
| 43 | Haute Loire | 51 | 68 |
| 44 | Loire Atlantique | 1294 | 1114 |
| 45 | Loiret | 927 | 870 |
| 46 | Lot | 50 | 97 |
| 47 | Lot Et Garonne | 71 | 143 |
| 48 | Lozere | 24 | 32 |
| 49 | Maine Et Loire | 240 | 328 |
| 50 | Manche | 101 | 196 |
| 51 | Marne | 152 | 240 |
| 52 | Haute Marne | 26 | 63 |
| 53 | Mayenne | 52 | 79 |
| 54 | Meurthe Et Moselle | 1687 | 1338 |
| 55 | Meuse | 38 | 85 |
| 56 | Morbihan | 232 | 441 |
| 57 | Moselle | 249 | 343 |
| 58 | Nievre | 41 | 86 |
| 59 | Nord | 1675 | 1261 |
| 60 | Oise | 142 | 198 |
| 61 | Orne | 40 | 80 |
| 62 | Pas De Calais | 479 | 547 |
| 63 | Puy De Dome | 249 | 331 |
| 64 | Pyrenees Atlantiques | 1026 | 948 |
| 65 | Hautes Pyrenees | 60 | 123 |
| 66 | Pyrenees Orientales | 98 | 239 |
| 67 | Bas Rhin | 399 | 421 |
| 68 | Haut Rhin | 905 | 541 |

Table 1: Number of serological samples for each department and age group *(continued)*

| Code | Depatment | Age group<br>Under 60 | Over 60 |
|---|---|---|---|
| 69 | Rhone | 2472 | 1767 |
| 70 | Haute Saone | 42 | 109 |
| 71 | Saone Et Loire | 137 | 276 |
| 72 | Sarthe | 553 | 481 |
| 73 | Savoie | 146 | 253 |
| 74 | Haute Savoie | 308 | 315 |
| 75 | Paris | 4799 | 2834 |
| 76 | Seine Maritime | 292 | 450 |
| 77 | Seine Et Marne | 338 | 452 |
| 78 | Yvelines | 670 | 701 |
| 79 | Deux Sevres | 93 | 155 |
| 80 | Somme | 98 | 154 |
| 81 | Tarn | 98 | 207 |
| 82 | Tarn Et Garonne | 53 | 101 |
| 83 | Var | 263 | 529 |
| 84 | Vaucluse | 115 | 305 |
| 85 | Vendee | 131 | 253 |
| 86 | Vienne | 785 | 673 |
| 87 | Haute Vienne | 117 | 188 |
| 88 | Vosges | 93 | 153 |
| 89 | Yonne | 331 | 338 |
| 90 | Territoire De Belfort | 35 | 72 |
| 91 | Essonne | 490 | 643 |
| 92 | Hauts De Seine | 1029 | 766 |
| 93 | Seine Saint Denis | 408 | 272 |
| 94 | Val De Marne | 650 | 581 |
| 95 | Val D Oise | 259 | 342 |

#### Departmental estimates and characteristics

##### Incidence, infection fatality rate (IFR) and infection hospitalization rate (IHR)

Table 2: Departmental incidence, infection fatality rate and infection hospitalization rate. Mean, q2.5, q97.5: mean of the posterior distribution and bounds of the credible interval.

| | | Incidence (%) | | | IFR (%) | | | IHR (%) | | |
|---|---|---|---|---|---|---|---|---|---|---|
| | | Mean | q2.5 | q97.5 | Mean | q2.5 | q97.5 | Mean | q2.5 | q97.5 |
| 1 | Ain | 7.14 | 5.98 | 8.44 | 0.77 | 0.63 | 0.94 | 1.85 | 1.52 | 2.22 |
| 2 | Aisne | 8.94 | 7.63 | 10.33 | 1.39 | 1.17 | 1.63 | 4.21 | 3.61 | 4.91 |
| 3 | Allier | 4.25 | 3.34 | 5.27 | 0.51 | 0.37 | 0.67 | 2.32 | 1.81 | 2.97 |
| 4 | Alpes De Haute Provence | 4.73 | 3.71 | 5.90 | 0.44 | 0.31 | 0.60 | 2.53 | 1.96 | 3.26 |
| 5 | Hautes Alpes | 5.11 | 4.01 | 6.39 | 0.43 | 0.29 | 0.59 | 2.54 | 1.95 | 3.25 |
| 6 | Alpes Maritimes | 4.97 | 4.05 | 5.97 | 0.57 | 0.46 | 0.70 | 2.34 | 1.91 | 2.88 |
| 7 | Ardeche | 7.76 | 6.51 | 9.15 | 1.33 | 1.09 | 1.60 | 3.33 | 2.78 | 3.98 |
| 8 | Ardennes | 6.43 | 5.22 | 7.75 | 0.76 | 0.60 | 0.96 | 2.79 | 2.26 | 3.46 |
| 9 | Ariege | 2.53 | 1.78 | 3.41 | 0.30 | 0.18 | 0.45 | 1.48 | 1.01 | 2.13 |
| 10 | Aube | 8.28 | 7.06 | 9.69 | 0.77 | 0.62 | 0.94 | 4.93 | 4.15 | 5.78 |
| 11 | Aude | 5.28 | 4.28 | 6.36 | 0.76 | 0.60 | 0.96 | 2.90 | 2.34 | 3.56 |
| 12 | Aveyron | 4.35 | 3.44 | 5.36 | 0.67 | 0.51 | 0.88 | 1.85 | 1.43 | 2.37 |
| 13 | Bouches Du Rhone | 5.99 | 5.19 | 6.84 | 0.71 | 0.61 | 0.83 | 4.73 | 4.11 | 5.44 |
| 14 | Calvados | 4.99 | 4.17 | 5.88 | 0.39 | 0.31 | 0.49 | 1.80 | 1.49 | 2.17 |
| 15 | Cantal | 3.39 | 2.55 | 4.33 | 0.39 | 0.25 | 0.55 | 1.84 | 1.34 | 2.48 |
| 16 | Charente | 3.41 | 2.73 | 4.16 | 0.44 | 0.32 | 0.58 | 1.19 | 0.91 | 1.53 |
| 17 | Charente Maritime | 3.78 | 2.99 | 4.66 | 0.47 | 0.36 | 0.61 | 1.59 | 1.24 | 2.01 |
| 18 | Cher | 6.10 | 4.99 | 7.32 | 1.10 | 0.88 | 1.35 | 2.21 | 1.79 | 2.72 |
| 19 | Correze | 4.54 | 3.61 | 5.55 | 0.68 | 0.51 | 0.89 | 2.53 | 1.99 | 3.21 |
| 20 | Corse | 5.47 | 4.42 | 6.63 | 0.51 | 0.39 | 0.66 | 2.73 | 2.20 | 3.37 |
| 21 | Cote D Or | 8.46 | 7.30 | 9.76 | 1.06 | 0.90 | 1.25 | 3.84 | 3.28 | 4.45 |
| 22 | Cotes D Armor | 3.54 | 2.85 | 4.32 | 0.46 | 0.35 | 0.60 | 1.70 | 1.34 | 2.14 |
| 23 | Creuse | 4.32 | 3.34 | 5.42 | 0.64 | 0.44 | 0.88 | 2.69 | 2.02 | 3.55 |
| 24 | Dordogne | 3.13 | 2.41 | 3.93 | 0.43 | 0.31 | 0.58 | 1.75 | 1.33 | 2.29 |
| 25 | Doubs | 8.69 | 7.42 | 10.12 | 1.25 | 1.05 | 1.48 | 2.25 | 1.90 | 2.66 |
| 26 | Drome | 6.92 | 5.84 | 8.12 | 0.76 | 0.63 | 0.92 | 2.67 | 2.24 | 3.17 |
| 27 | Eure | 5.83 | 4.76 | 6.93 | 0.84 | 0.68 | 1.03 | 1.42 | 1.16 | 1.75 |
| 28 | Eure Et Loir | 7.49 | 6.37 | 8.71 | 0.99 | 0.83 | 1.18 | 3.15 | 2.65 | 3.71 |

Table 2: Departmental incidence, infection fatality rate and infection hospitalization rate. Mean, q2.5, q97.5: mean of the posterior distribution and bounds of the credible interval. *(continued)*

| | | Incidence (%) | | | IFR (%) | | | IHR (%) | | |
|---|---|---|---|---|---|---|---|---|---|---|
| | | Mean | q2.5 | q97.5 | Mean | q2.5 | q97.5 | Mean | q2.5 | q97.5 |
| 29 | Finistere | 3.04 | 2.32 | 3.86 | 0.32 | 0.24 | 0.43 | 1.12 | 0.84 | 1.46 |
| 30 | Gard | 4.35 | 3.59 | 5.16 | 0.73 | 0.59 | 0.90 | 1.84 | 1.51 | 2.23 |
| 31 | Haute Garonne | 3.94 | 3.32 | 4.58 | 0.27 | 0.21 | 0.33 | 1.77 | 1.49 | 2.11 |
| 32 | Gers | 4.71 | 3.71 | 5.82 | 0.79 | 0.59 | 1.04 | 2.02 | 1.55 | 2.59 |
| 33 | Gironde | 4.74 | 4.07 | 5.43 | 0.42 | 0.35 | 0.50 | 2.27 | 1.95 | 2.65 |
| 34 | Herault | 4.77 | 3.91 | 5.68 | 0.52 | 0.42 | 0.65 | 2.01 | 1.65 | 2.44 |
| 35 | Ille Et Vilaine | 4.58 | 3.88 | 5.32 | 0.38 | 0.30 | 0.47 | 1.63 | 1.37 | 1.96 |
| 36 | Indre | 7.10 | 5.80 | 8.50 | 1.48 | 1.20 | 1.83 | 2.32 | 1.87 | 2.87 |
| 37 | Indre Et Loire | 5.34 | 4.59 | 6.13 | 0.59 | 0.48 | 0.71 | 2.01 | 1.70 | 2.36 |
| 38 | Isere | 5.60 | 4.74 | 6.50 | 0.64 | 0.53 | 0.76 | 1.78 | 1.50 | 2.10 |
| 39 | Jura | 7.36 | 6.17 | 8.70 | 0.90 | 0.72 | 1.10 | 2.72 | 2.24 | 3.28 |
| 40 | Landes | 2.89 | 2.15 | 3.74 | 0.36 | 0.25 | 0.49 | 1.06 | 0.77 | 1.43 |
| 41 | Loir Et Cher | 5.92 | 4.94 | 6.97 | 0.81 | 0.65 | 1.00 | 2.80 | 2.32 | 3.38 |
| 42 | Loire | 7.33 | 6.24 | 8.49 | 1.17 | 0.99 | 1.38 | 3.24 | 2.75 | 3.79 |
| 43 | Haute Loire | 4.22 | 3.31 | 5.27 | 0.44 | 0.31 | 0.60 | 2.13 | 1.64 | 2.76 |
| 44 | Loire Atlantique | 5.55 | 4.74 | 6.39 | 0.50 | 0.42 | 0.60 | 1.55 | 1.32 | 1.82 |
| 45 | Loiret | 5.95 | 5.11 | 6.82 | 0.55 | 0.45 | 0.66 | 2.80 | 2.39 | 3.28 |
| 46 | Lot | 4.34 | 3.39 | 5.43 | 0.56 | 0.41 | 0.76 | 3.19 | 2.47 | 4.10 |
| 47 | Lot Et Garonne | 2.75 | 2.05 | 3.52 | 0.34 | 0.23 | 0.48 | 1.40 | 1.03 | 1.89 |
| 48 | Lozere | 3.20 | 2.28 | 4.22 | 0.30 | 0.18 | 0.47 | 1.31 | 0.87 | 1.90 |
| 49 | Maine Et Loire | 5.88 | 4.91 | 6.89 | 0.67 | 0.56 | 0.82 | 2.26 | 1.89 | 2.69 |
| 50 | Manche | 4.21 | 3.34 | 5.20 | 0.48 | 0.36 | 0.62 | 1.54 | 1.20 | 1.96 |
| 51 | Marne | 9.08 | 7.87 | 10.42 | 1.16 | 0.98 | 1.35 | 3.70 | 3.19 | 4.26 |
| 52 | Haute Marne | 9.00 | 7.69 | 10.57 | 1.39 | 1.14 | 1.67 | 3.83 | 3.19 | 4.54 |
| 53 | Mayenne | 5.24 | 4.18 | 6.43 | 0.52 | 0.39 | 0.68 | 2.00 | 1.58 | 2.53 |
| 54 | Meurthe Et Moselle | 8.02 | 7.17 | 8.94 | 0.99 | 0.86 | 1.13 | 3.93 | 3.49 | 4.42 |
| 55 | Meuse | 10.06 | 8.70 | 11.62 | 1.29 | 1.06 | 1.54 | 5.20 | 4.44 | 6.01 |
| 56 | Morbihan | 4.60 | 3.70 | 5.60 | 0.47 | 0.36 | 0.59 | 1.86 | 1.50 | 2.32 |
| 57 | Moselle | 10.48 | 9.16 | 12.00 | 1.41 | 1.22 | 1.61 | 4.23 | 3.67 | 4.83 |
| 58 | Nievre | 4.68 | 3.62 | 5.87 | 0.86 | 0.64 | 1.14 | 1.87 | 1.41 | 2.45 |
| 59 | Nord | 6.00 | 5.18 | 6.83 | 0.80 | 0.69 | 0.94 | 3.02 | 2.63 | 3.49 |
| 60 | Oise | 8.93 | 7.66 | 10.31 | 1.21 | 1.03 | 1.42 | 3.14 | 2.68 | 3.66 |

Table 2: Departmental incidence, infection fatality rate and infection hospitalization rate. Mean, q2.5, q97.5: mean of the posterior distribution and bounds of the credible interval. *(continued)*

| | | Incidence (%) | | | IFR (%) | | | IHR (%) | | |
|---|---|---|---|---|---|---|---|---|---|---|
| | | Mean | q2.5 | q97.5 | Mean | q2.5 | q97.5 | Mean | q2.5 | q97.5 |
| 61 | Orne | 5.38 | 4.36 | 6.49 | 0.60 | 0.46 | 0.77 | 2.73 | 2.21 | 3.38 |
| 62 | Pas De Calais | 6.09 | 5.09 | 7.14 | 0.79 | 0.66 | 0.95 | 2.97 | 2.51 | 3.53 |
| 63 | Puy De Dome | 4.02 | 3.22 | 4.90 | 0.46 | 0.36 | 0.59 | 1.30 | 1.02 | 1.62 |
| 64 | Pyrenees Atlantiques | 3.86 | 3.18 | 4.61 | 0.36 | 0.27 | 0.46 | 1.31 | 1.06 | 1.62 |
| 65 | Hautes Pyrenees | 5.07 | 4.01 | 6.25 | 0.78 | 0.60 | 1.00 | 2.15 | 1.67 | 2.72 |
| 66 | Pyrenees Orientales | 3.66 | 2.78 | 4.62 | 0.42 | 0.31 | 0.57 | 2.68 | 2.06 | 3.49 |
| 67 | Bas Rhin | 10.67 | 9.34 | 12.26 | 1.17 | 1.00 | 1.34 | 3.78 | 3.27 | 4.31 |
| 68 | Haut Rhin | 13.60 | 12.12 | 15.22 | 1.97 | 1.74 | 2.22 | 5.20 | 4.60 | 5.84 |
| 69 | Rhone | 7.57 | 6.73 | 8.42 | 1.14 | 1.01 | 1.29 | 4.11 | 3.68 | 4.62 |
| 70 | Haute Saone | 8.42 | 7.13 | 9.89 | 1.27 | 1.04 | 1.53 | 2.50 | 2.05 | 2.99 |
| 71 | Saone Et Loire | 6.90 | 5.84 | 8.03 | 1.05 | 0.87 | 1.25 | 3.85 | 3.26 | 4.54 |
| 72 | Sarthe | 5.31 | 4.44 | 6.20 | 0.64 | 0.51 | 0.78 | 2.42 | 2.02 | 2.90 |
| 73 | Savoie | 6.43 | 5.31 | 7.67 | 0.53 | 0.42 | 0.66 | 2.33 | 1.92 | 2.83 |
| 74 | Haute Savoie | 7.49 | 6.28 | 8.82 | 0.77 | 0.64 | 0.93 | 2.29 | 1.91 | 2.73 |
| 75 | Paris | 10.12 | 9.34 | 10.97 | 1.10 | 1.00 | 1.20 | 5.15 | 4.73 | 5.60 |
| 76 | Seine Maritime | 5.17 | 4.24 | 6.13 | 0.59 | 0.48 | 0.72 | 2.24 | 1.86 | 2.73 |
| 77 | Seine Et Marne | 9.82 | 8.62 | 11.17 | 1.04 | 0.90 | 1.19 | 3.60 | 3.15 | 4.10 |
| 78 | Yvelines | 9.43 | 8.28 | 10.68 | 1.13 | 0.99 | 1.30 | 3.19 | 2.79 | 3.62 |
| 79 | Deux Sevres | 3.51 | 2.71 | 4.39 | 0.42 | 0.30 | 0.57 | 1.20 | 0.90 | 1.58 |
| 80 | Somme | 8.03 | 6.82 | 9.34 | 1.24 | 1.05 | 1.48 | 3.16 | 2.67 | 3.72 |
| 81 | Tarn | 3.76 | 2.92 | 4.68 | 0.46 | 0.34 | 0.61 | 1.39 | 1.06 | 1.80 |
| 82 | Tarn Et Garonne | 2.96 | 2.20 | 3.80 | 0.36 | 0.24 | 0.51 | 1.02 | 0.72 | 1.42 |
| 83 | Var | 5.12 | 4.24 | 6.07 | 0.63 | 0.51 | 0.77 | 2.58 | 2.14 | 3.10 |
| 84 | Vaucluse | 4.65 | 3.76 | 5.62 | 0.40 | 0.30 | 0.51 | 1.79 | 1.44 | 2.22 |
| 85 | Vendee | 4.19 | 3.35 | 5.13 | 0.49 | 0.38 | 0.62 | 1.61 | 1.28 | 2.02 |
| 86 | Vienne | 4.47 | 3.72 | 5.28 | 0.42 | 0.32 | 0.54 | 1.38 | 1.12 | 1.70 |
| 87 | Haute Vienne | 3.75 | 2.85 | 4.73 | 0.47 | 0.34 | 0.64 | 1.61 | 1.21 | 2.13 |
| 88 | Vosges | 10.85 | 9.35 | 12.68 | 1.63 | 1.37 | 1.91 | 3.28 | 2.77 | 3.83 |
| 89 | Yonne | 6.66 | 5.64 | 7.77 | 1.05 | 0.87 | 1.28 | 2.82 | 2.35 | 3.36 |
| 90 | Territoire De Belfort | 11.49 | 10.05 | 13.25 | 1.37 | 1.12 | 1.63 | 5.92 | 5.09 | 6.79 |
| 91 | Essonne | 9.70 | 8.52 | 10.98 | 1.20 | 1.04 | 1.37 | 3.66 | 3.21 | 4.16 |
| 92 | Hauts De Seine | 10.98 | 9.85 | 12.21 | 1.18 | 1.05 | 1.33 | 4.82 | 4.30 | 5.37 |

Table 2: Departmental incidence, infection fatality rate and infection hospitalization rate. Mean, q2.5, q97.5: mean of the posterior distribution and bounds of the credible interval. *(continued)*

| | | Incidence (%) | | | IFR (%) | | | IHR (%) | | |
|---|---|---|---|---|---|---|---|---|---|---|
| | | Mean | q2.5 | q97.5 | Mean | q2.5 | q97.5 | Mean | q2.5 | q97.5 |
| 93 | Seine Saint Denis | 10.76 | 9.31 | 12.36 | 1.23 | 1.06 | 1.42 | 4.52 | 3.92 | 5.21 |
| 94 | Val De Marne | 11.75 | 10.48 | 13.15 | 1.27 | 1.12 | 1.43 | 5.18 | 4.60 | 5.80 |
| 95 | Val D Oise | 10.44 | 9.10 | 11.94 | 1.41 | 1.22 | 1.62 | 3.99 | 3.46 | 4.57 |

#### Departmental characteristics (data)

Table 3: Departmental characteristics (data). Pop.: Size of the adult population. ICU beds: Number of intensive care beds per 1,000 inhabitant. Diabetes: Prevalence of diabetes. Pop. 60: Proportion of the population over 60. Deaths: Number of deaths attributed to COVID-19. Hospit.: Number of hospitalization attributed to COVID-19.

| Code | Depatment | Pop. | ICU beds | Diabetes | Pop. 60 | Deaths | Hospit. |
|---|---|---|---|---|---|---|---|
| 1 | Ain | 487636 | 0.13 | 4.33 % | 32 % | 268 | 635 |
| 2 | Aisne | 398119 | 0.31 | 6.72 % | 37 % | 494 | 1492 |
| 3 | Allier | 266989 | 0.29 | 6.85 % | 44 % | 53 | 261 |
| 4 | Alpes De Haute Provence | 130747 | 0.12 | 5.42 % | 43 % | 23 | 158 |
| 5 | Hautes Alpes | 110923 | 0.24 | 4.32 % | 42 % | 21 | 146 |
| 6 | Alpes Maritimes | 865545 | 0.52 | 5.42 % | 39 % | 243 | 997 |
| 7 | Ardeche | 257083 | 0.20 | 5.43 % | 42 % | 271 | 662 |
| 8 | Ardennes | 206499 | 0.22 | 6.72 % | 38 % | 98 | 367 |
| 9 | Ariege | 122112 | 0.18 | 5.6 % | 43 % | 5 | 45 |
| 10 | Aube | 235127 | 0.29 | 5.95 % | 37 % | 143 | 963 |
| 11 | Aude | 295187 | 0.24 | 6.34 % | 43 % | 119 | 450 |
| 12 | Aveyron | 223024 | 0.22 | 5.08 % | 44 % | 69 | 177 |
| 13 | Bouches Du Rhone | 1567212 | 0.60 | 5.78 % | 35 % | 668 | 4428 |
| 14 | Calvados | 533685 | 0.50 | 4.88 % | 37 % | 100 | 474 |
| 15 | Cantal | 116890 | 0.38 | 5.76 % | 45 % | 12 | 72 |
| 16 | Charente | 277989 | 0.23 | 5.72 % | 42 % | 40 | 108 |
| 17 | Charente Maritime | 522684 | 0.22 | 5.61 % | 45 % | 91 | 309 |
| 18 | Cher | 237185 | 0.30 | 6.68 % | 42 % | 162 | 314 |
| 19 | Correze | 191475 | 0.37 | 6.11 % | 44 % | 60 | 219 |
| 20 | Corse | 274777 | 0.23 | 5.04 % | 38 % | 74 | 409 |
| 21 | Cote D Or | 413687 | 0.63 | 5.07 % | 36 % | 371 | 1339 |
| 22 | Cotes D Armor | 468624 | 0.18 | 4.29 % | 43 % | 78 | 280 |
| 23 | Creuse | 94918 | 0.29 | 7.37 % | 48 % | 24 | 111 |
| 24 | Dordogne | 334270 | 0.21 | 6.29 % | 46 % | 42 | 181 |
| 25 | Doubs | 410960 | 0.67 | 4.96 % | 34 % | 448 | 791 |
| 26 | Drome | 393336 | 0.25 | 5.26 % | 38 % | 206 | 722 |
| 27 | Eure | 446217 | 0.11 | 5.3 % | 35 % | 220 | 361 |
| 28 | Eure Et Loir | 322625 | 0.26 | 5.62 % | 36 % | 240 | 757 |
| 29 | Finistere | 713244 | 0.32 | 3.89 % | 39 % | 69 | 236 |
| 30 | Gard | 579457 | 0.31 | 5.69 % | 39 % | 188 | 458 |
| 31 | Haute Garonne | 1081415 | 0.62 | 3.93 % | 29 % | 108 | 748 |

Table 3: Departmental characteristics (data). Pop.: Size of the adult population. ICU beds: Number of intensive care beds per 1,000 inhabitant. Diabetes: Prevalence of diabetes. Pop. 60: Proportion of the population over 60. Deaths: Number of deaths attributed to COVID-19. Hospit.: Number of hospitalization attributed to COVID-19. *(continued)*

| Code | Depatment | Pop. | ICU beds | Diabetes | Pop. 60 | Deaths | Hospit. |
|---|---|---|---|---|---|---|---|
| 32 | Gers | 152962 | 0.13 | 5.41 % | 45 % | 62 | 144 |
| 33 | Gironde | 1259223 | 0.47 | 4.53 % | 33 % | 249 | 1349 |
| 34 | Herault | 918558 | 0.57 | 5.15 % | 36 % | 227 | 870 |
| 35 | Ille Et Vilaine | 815134 | 0.38 | 3.07 % | 32 % | 141 | 607 |
| 36 | Indre | 174947 | 0.23 | 6.95 % | 45 % | 191 | 281 |
| 37 | Indre Et Loire | 469886 | 0.59 | 5.06 % | 37 % | 147 | 498 |
| 38 | Isere | 953296 | 0.27 | 4.75 % | 33 % | 340 | 940 |
| 39 | Jura | 200336 | 0.14 | 5.3 % | 40 % | 132 | 399 |
| 40 | Landes | 330021 | 0.18 | 5.62 % | 42 % | 31 | 96 |
| 41 | Loir Et Cher | 254990 | 0.22 | 6.21 % | 42 % | 122 | 421 |
| 42 | Loire | 581757 | 0.38 | 5.53 % | 38 % | 503 | 1373 |
| 43 | Haute Loire | 176577 | 0.18 | 5.36 % | 41 % | 30 | 159 |
| 44 | Loire Atlantique | 1082428 | 0.34 | 3.61 % | 33 % | 299 | 923 |
| 45 | Loiret | 510770 | 0.47 | 5.54 % | 35 % | 161 | 847 |
| 46 | Lot | 141616 | 0.16 | 5.76 % | 47 % | 34 | 202 |
| 47 | Lot Et Garonne | 260024 | 0.25 | 5.75 % | 43 % | 21 | 97 |
| 48 | Lozere | 60607 | 0.10 | 4.76 % | 41 % | 2 | 24 |
| 49 | Maine Et Loire | 613033 | 0.39 | 4.29 % | 36 % | 244 | 808 |
| 50 | Manche | 386850 | 0.23 | 4.89 % | 41 % | 77 | 247 |
| 51 | Marne | 430280 | 0.39 | 5.78 % | 34 % | 449 | 1436 |
| 52 | Haute Marne | 135508 | 0.37 | 6.72 % | 42 % | 170 | 463 |
| 53 | Mayenne | 230099 | 0.13 | 3.9 % | 38 % | 63 | 240 |
| 54 | Meurthe Et Moselle | 562144 | 0.62 | 5.54 % | 34 % | 445 | 1769 |
| 55 | Meuse | 141967 | 0.21 | 6.12 % | 40 % | 181 | 745 |
| 56 | Morbihan | 594042 | 0.21 | 4.21 % | 41 % | 126 | 505 |
| 57 | Moselle | 814411 | 0.29 | 6.18 % | 35 % | 1194 | 3589 |
| 58 | Nievre | 163421 | 0.29 | 7.94 % | 47 % | 66 | 137 |
| 59 | Nord | 1926717 | 0.50 | 5.76 % | 31 % | 928 | 3478 |
| 60 | Oise | 610459 | 0.21 | 5.5 % | 32 % | 661 | 1699 |
| 61 | Orne | 217351 | 0.23 | 5.82 % | 43 % | 68 | 319 |
| 62 | Pas De Calais | 1089833 | 0.36 | 6.36 % | 35 % | 522 | 1959 |
| 63 | Puy De Dome | 512938 | 0.44 | 5.14 % | 37 % | 95 | 260 |
| 64 | Pyrenees Atlantiques | 542011 | 0.34 | 4.74 % | 40 % | 72 | 269 |

Table 3: Departmental characteristics (data). Pop.: Size of the adult population. ICU beds: Number of intensive care beds per 1,000 inhabitant. Diabetes: Prevalence of diabetes. Pop. 60: Proportion of the population over 60. Deaths: Number of deaths attributed to COVID-19. Hospit.: Number of hospitalization attributed to COVID-19. *(continued)*

| Code | Depatment | Pop. | ICU beds | Diabetes | Pop. 60 | Deaths | Hospit. |
|---|---|---|---|---|---|---|---|
| 65 | Hautes Pyrenees | 183708 | 0.39 | 5.6 % | 43 % | 76 | 197 |
| 66 | Pyrenees Orientales | 377631 | 0.28 | 5.93 % | 42 % | 55 | 370 |
| 67 | Bas Rhin | 886074 | 0.52 | 5.71 % | 33 % | 1092 | 3556 |
| 68 | Haut Rhin | 589919 | 0.42 | 5.74 % | 35 % | 1573 | 4153 |
| 69 | Rhone | 1407653 | 0.51 | 4.55 % | 30 % | 1218 | 4379 |
| 70 | Haute Saone | 180375 | 0.27 | 6.04 % | 39 % | 194 | 372 |
| 71 | Saone Et Loire | 431699 | 0.26 | 6.44 % | 43 % | 312 | 1144 |
| 72 | Sarthe | 429348 | 0.22 | 5.25 % | 38 % | 145 | 549 |
| 73 | Savoie | 338016 | 0.25 | 4.22 % | 35 % | 111 | 502 |
| 74 | Haute Savoie | 628526 | 0.26 | 3.67 % | 30 % | 364 | 1067 |
| 75 | Paris | 1740626 | 0.83 | 4.04 % | 28 % | 1929 | 9071 |
| 76 | Seine Maritime | 949467 | 0.46 | 5.54 % | 35 % | 286 | 1091 |
| 77 | Seine Et Marne | 1037205 | 0.25 | 4.96 % | 28 % | 1050 | 3651 |
| 78 | Yvelines | 1062664 | 0.28 | 4.23 % | 30 % | 1131 | 3179 |
| 79 | Deux Sevres | 288272 | 0.17 | 5.32 % | 40 % | 41 | 117 |
| 80 | Somme | 431476 | 0.57 | 6.2 % | 35 % | 432 | 1085 |
| 81 | Tarn | 305490 | 0.33 | 5.12 % | 42 % | 53 | 155 |
| 82 | Tarn Et Garonne | 198708 | 0.28 | 5.21 % | 38 % | 20 | 55 |
| 83 | Var | 859946 | 0.25 | 5.69 % | 42 % | 275 | 1127 |
| 84 | Vaucluse | 429524 | 0.31 | 5.87 % | 38 % | 73 | 352 |
| 85 | Vendee | 533161 | 0.21 | 5.08 % | 42 % | 108 | 355 |
| 86 | Vienne | 337739 | 0.49 | 4.87 % | 37 % | 61 | 203 |
| 87 | Haute Vienne | 293360 | 0.75 | 5.96 % | 40 % | 50 | 171 |
| 88 | Vosges | 283485 | 0.21 | 5.95 % | 41 % | 500 | 999 |
| 89 | Yonne | 258366 | 0.26 | 6.56 % | 41 % | 183 | 481 |
| 90 | Territoire De Belfort | 106390 | 0.54 | 5.31 % | 35 % | 163 | 728 |
| 91 | Essonne | 952194 | 0.33 | 4.62 % | 29 % | 1104 | 3367 |
| 92 | Hauts De Seine | 1224112 | 0.54 | 3.99 % | 27 % | 1584 | 6454 |
| 93 | Seine Saint Denis | 1174096 | 0.30 | 5.87 % | 24 % | 1542 | 5685 |
| 94 | Val De Marne | 1048359 | 0.48 | 4.74 % | 28 % | 1554 | 6357 |
| 95 | Val D Oise | 894870 | 0.20 | 5.32 % | 27 % | 1312 | 3710 |

### Complementary results

#### Univariate analysis

In univariate analysis, a departmental incidence of 3% was associated with an IFR of 0.41% (95% CI: 0.33-0.49%), and an incidence of 9% was associated with an IFR of 1.09% (95% CI: 0.94-1.28%). The absolute difference was 0.69% (95% CI: 0.52-0.89%).
An incidence of 3% was associated with an IHR of 1.57% (95% CI: 1.27-1.89%), and an incidence of 9% was associated with an IHR of 3.55% (95% CI: 3.08-4.08%). The absolute difference was 1.98% (95% CI: 1.43-2.60%).

#### Role of the confounders (association between incidence, IFR and IHR)

Table 4: Posterior mean and 95% CI of the parameters of equation 14 (main manuscript, Methods section). Covariates are considered at the departmental scale. Age: Proportion of the population over 60. Diabetes: Prevalence of diabetes. Beds: Number of intensive care beds per 1,000 inhabitant.

| Outcome | Covariate | Parameter | mean | q2.5 | q97.5 |
|---|---|---|---|---|---|
| IFR | Age | $c_{age}$ | 0.452 | -2.790 | 3.869 |
| IFR | Beds | $c_{beds}$ | -0.054 | -0.710 | 0.619 |
| IFR | Diabetes | $c_{diab}$ | 16.402 | 1.255 | 31.260 |
| IHR | Age | $d_{age}$ | 0.061 | -3.584 | 3.880 |
| IHR | Beds | $d_{beds}$ | 0.501 | -0.230 | 1.244 |
| IHR | Diabetes | $d_{diab}$ | 12.381 | -4.701 | 28.482 |

#### Computation of expectations

##### Expected proportion of persons over 60 in the infected given incidence in the 20-59 in a department with the same age structure as metropolitan France

On the logit scale, the distribution of incidence in persons over 60 (logit $U_1$) is normal given an incidence $u_0$ in the 20-59 (following equation 7 of the article):

$$\text{logit } U_1 | u_0 \sim N(\text{logit } u_0 + \mu_{\text{age}} + (\text{logit } u_0) \times b_{\text{age}}, \sigma_{\text{age}})$$

Thus, the expectation of logit $U_1$ given $u_0$ is:

$$E\left[\text{logit } U_1 | u_0\right] = \text{logit } u_0 + \mu_{\text{age}} + (\text{logit } u_0) \times b_{\text{age}}$$

As the logistic transformation is non-linear, in general the expectation of the transformed variable is not the transformation of the expectation:

$$E\left[U_1 | u_0\right] \neq \text{logit}^{-1} E\left[\text{logit } U_1 | u_0\right] \qquad \text{(in general)}$$

To overcome this limitation, the expectation $E\left[U_1|u_0\right]$ was approximated for each MCMC iteration by drawing 10,000 random values logit $u_1^*(u_0)$ from $P(\text{logit } U_1|u_0)$, then for each of these values, applying the logistic transformation and computing $\text{age}^*_{\text{infected}}(u_0)$, the proportion of persons over 60 among those infected given $u_0$ for a department with the same age structure as metropolitan France ($\text{age}_{\text{pop, France}}$):

$$\text{age}^*_{\text{infected}}(u_0) = \frac{u_1^*(u_0) \times \text{age}_{\text{pop, France}}}{u_1^*(u_0) \times \text{age}_{\text{pop, France}} + u_0 \times (1 - \text{age}_{\text{pop, France}})}$$

The 10,000 $\text{age}^*_{\text{infected}}(u_0)$ were averaged to obtain $E\left[U_1|u_0\right]$.

##### Expected causal effect of incidence on IFR and IHR

The linear predictor of IFR given incidence $x$ and given the covariates has a normal distribution (following equation 14 of the manuscript):

$$g_{\text{IFR}} | x, \text{diab}, \text{age}_{\text{pop}}, \text{beds} \sim N(m_{\text{IFR}} + x \times c_p + \text{diab} \times c_{\text{diab}} + \text{age}_{\text{pop}} \times c_{\text{age}} + \text{beds} \times c_{\text{beds}}, \sigma_{\text{IFR}})$$

Thus, the conditional expectation of this linear predictor is:

$$E\left[g_{\text{IFR}} | x, \text{diab}, \text{age}_{\text{pop}}, \text{beds}\right] = m_{\text{IFR}} + x \times c_p + \text{diab} \times c_{\text{diab}} + \text{age}_{\text{pop}} \times c_{\text{age}} + \text{beds} \times c_{\text{beds}}$$

The conditional expectation after logistic transformation was computed according to the same sampling procedure as for the proportion of persons over 60 among the infected, providing $E\left[\text{IFR}|x, \text{diab}, \text{age}_{\text{pop}}, \text{beds}\right]$ for any $x$, diab, $\text{age}_{\text{pop}}$, and beds.

Following equation 15 of the manuscript, using the linearity of expection, and using $j$ as an index for departments,

$$E\left[\text{IFR}|do(x)\right] = \frac{1}{95}\sum_{j=1}^{95} E\left[\text{IFR}|x, \text{diab}_j, \text{age}_{\text{pop},j}, \text{beds}_j\right]$$

The same procedure was used to compute the expected adjusted IHR given incidence.

#### Posterior predictive checks

Synthetic serological data, hospitalization counts, and death counts were drawn from the posterior predictive distribution to:

- Check the model versus the actual data
- Illustrate the importance of the Bayesian statistical framework in this study

The number of synthetic serological samples in each department is identical to the one in the actual data.

Figure 2, Figure 3, and Figure 4 consist in the comparison of the actual data against three randomly chosen HMC iterations in plots mimicking those presented in the article (but solely based on data). Naive linear regressions (naive in that they consider each department as a data point) were performed to show the linear relation between the two variables of each plot.

In Figure 2, Figure 3, and Figure 4, sampling variation leads to a spurious negative association between the quantity represented on the x-axis and the quantity represented on the y-axis. Indeed, the quantity represented on the x-axis is in the denominator of the quantity on the y-axis. Thus, when the quantity represented on the x-axis varies, the quantity represented on the y-axis tends to vary in the opposite direction. In the article, accounting for the variability of the latent variables used in the regressions highlighted a positive relationship between the variables, as observed below for the departments with the most serological data (the large points in Figure 2, Figure 3, and Figure 4).

Figure 5 shows that the value of the regression slope based on the actual data falls between the 2.5% and 97.5% quantiles of the distribution of regression slopes based on 1,000 posterior predictive draws, which satisfies the posterior predictive checks.

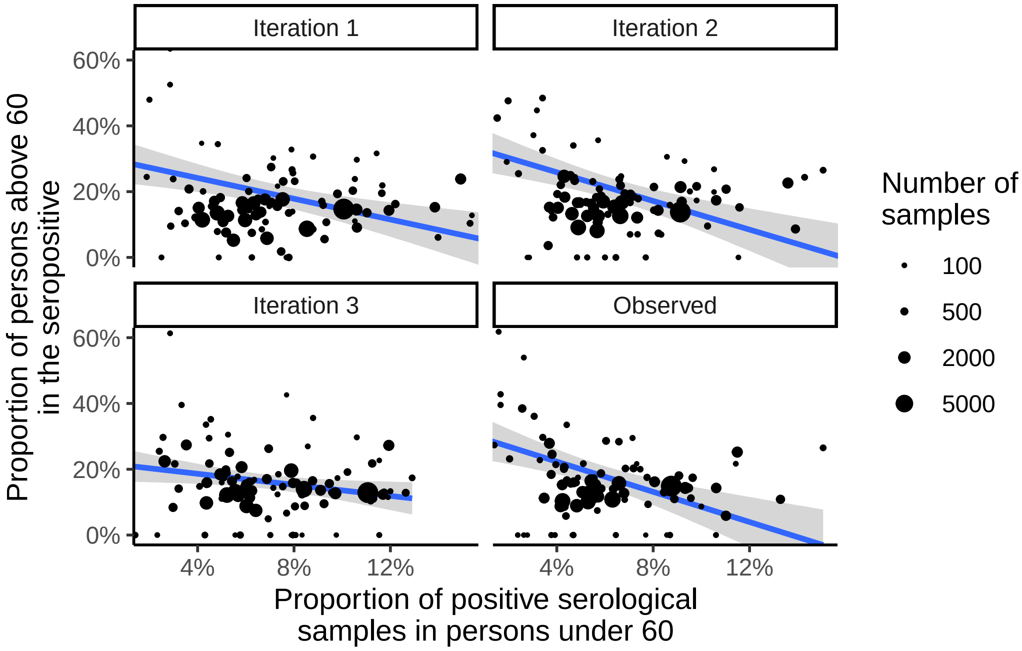

Figure 2: Posterior predictive check for the association between the proportion of positive serological samples in persons under 60 and the proportion of persons above 60 in the seropositive (mimicks Figure 3 of the article). The size of the points is proportional to the number of serological samples in the deparment. A naive linear regression (one department being one observation) illustrates this association. The y-axis is cut at 60% for readability (some points may not appear).

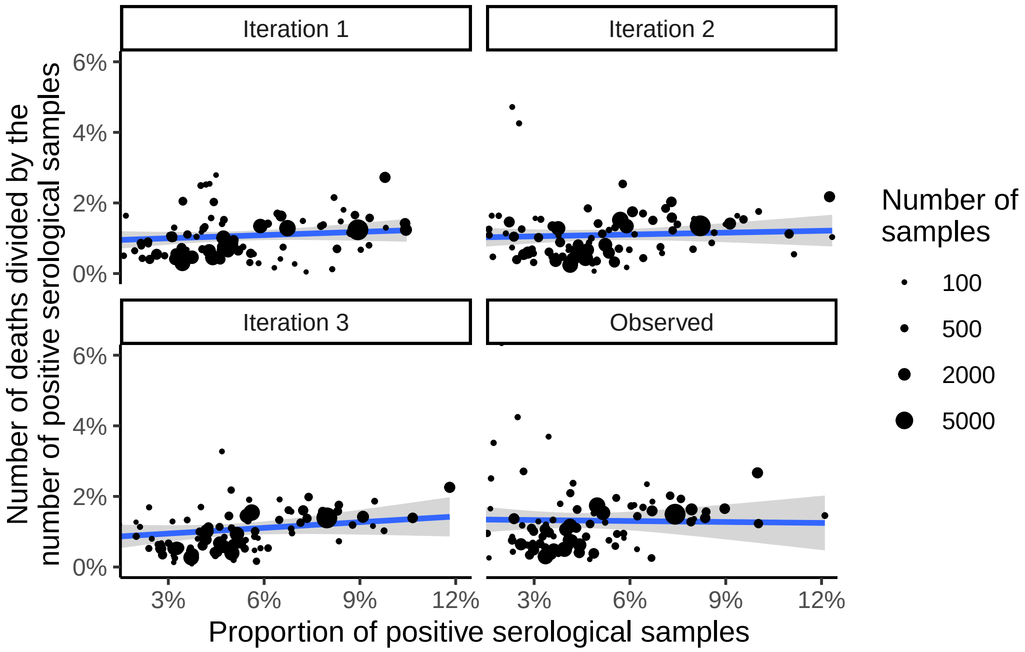

Figure 3: Posterior predictive check for the association between the proportion of positive serological samples and the number of COVID-19-related deaths divided by the number of positive serological samples. The size of the points is proportional to the number of serological samples in the deparment. A naive linear regression (one department being one observation) illustrates this association. The y-axis is cut at 6% for readability (some points may not appear).

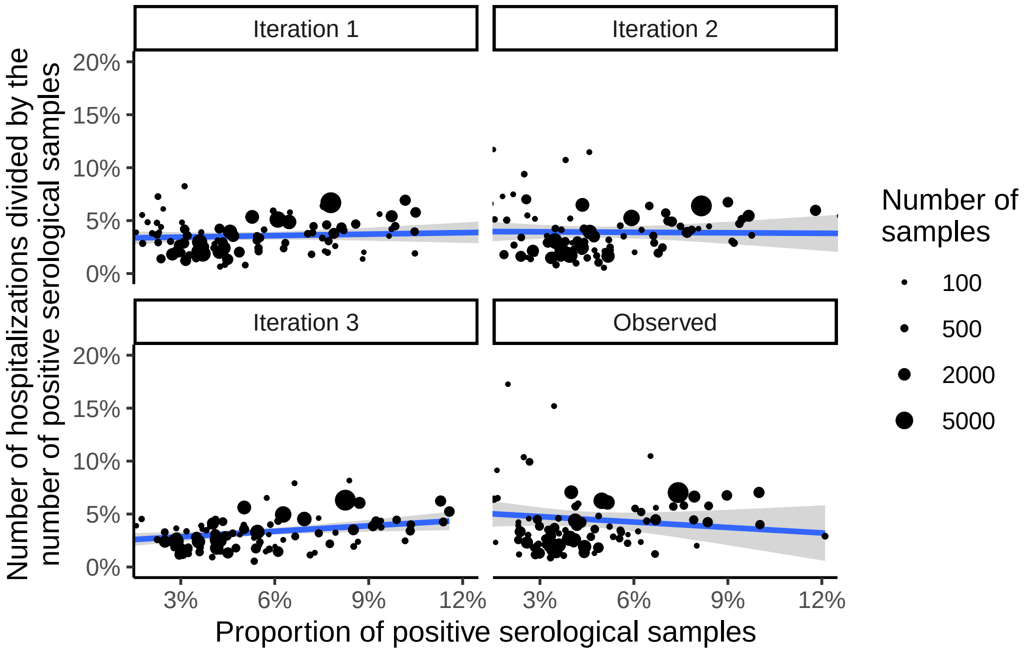

Figure 4: Posterior predictive check for the association between the proportion of positive serological samples and the number of COVID-19-related hospitalizations divided by the number of positive serological samples. The size of the points is proportional to the number of serological samples in the deparment. A naive linear regression (one department being one observation) illustrates this association. The y-axis is cut at 20% for readability (some points may not appear).

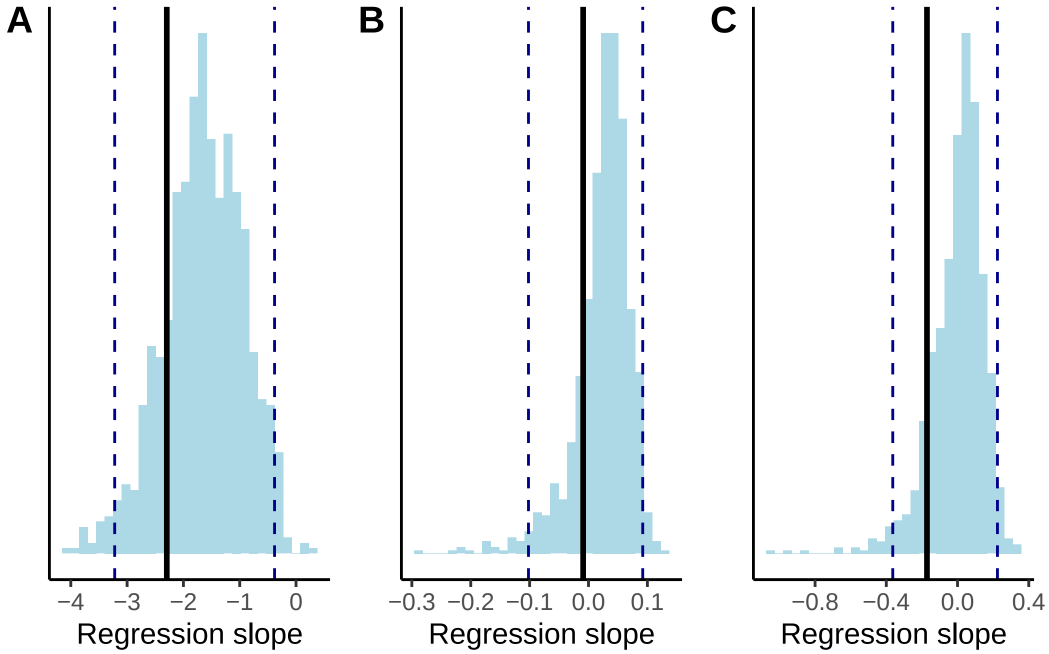

Figure 5: Quantitative posterior predictive check for: (A) the association between the proportion of positive serological samples in persons under 60 and the proportion of persons above 60 in the seropositive, (B) the association between the proportion of positive serological samples and the number of COVID-19-related deaths divided by the number of positive serological samples, and (C) the association between the proportion of positive serological samples and the number of COVID-19-related hospitalizations divided by the number of positive serological samples. The blue histogram represents the distribution of 1,000 regression slopes based on the posterior predictive draws (quantiles 2.5% and 97.5% are represented with the two blue dashed vertical lines). The black line corresponds to the regression slope based on the actual data.

#### Stan code

```
functions {
  // Custom distribution function icar_normal_lpdf for an ICAR random
  // variable phi (see Morris et al., 2019)
  real icar_normal_lpdf(
    vector phi, int N_dept, array[] int node1, array[] int node2
  ) {
    return -0.5 * dot_self(phi[node1] - phi[node2])
    + normal_lpdf(sum(phi) | 0, 0.001 * N_dept);
  }
}

data {

  int<lower=0> N_edges;
  array[N_edges] int<lower=1> node1;
  array[N_edges] int<lower=1> node2;

  int<lower=0> N_age;
  int<lower=0> N_dept;

  int<lower=0> N_pos_incid;
  array[N_pos_incid] int<lower=0> age_pos_incid;
  array[N_pos_incid] int<lower=0> dept_pos_incid;

  int<lower=0> N_sens_sapris;
  int<lower=0> y_sens_sapris;

  array[N_age, N_dept] int<lower=0> mat_N_age_dept;
  array[N_age, N_dept] int<lower=0> mat_y_age_dept;

  vector<lower=0, upper=1>[N_dept] prop_dept;
  vector<lower=0, upper=1>[N_age] prop_age;
  matrix<lower=0, upper=1>[N_dept, N_age] mat_dept_age;
  matrix<lower=0, upper=1>[N_age, N_dept] mat_age_dept;

  int<lower=0> N_grid_Y;
  vector<lower=0, upper=1>[N_grid_Y] grid_Y;
```

```
  vector<lower=0>[N_dept] pop_dept;
  array[N_dept] int<lower=0> hospit_dept;
  array[N_dept] int<lower=0> deces_dept;

  vector<lower=0, upper=1>[N_dept] age_pop;
  vector<lower=0, upper=1>[N_dept] beds;
  vector<lower=0, upper=1>[N_dept] diabetes;

  int<lower=0> N_draws;

}

parameters {

  vector[N_dept] phi_raw; // Corresponds to the parameter phi of the manuscript
  real<lower=0> sigma_phi;

  real<lower=0, upper=1> sens;
  real<lower=0, upper=1> spe;

  real mu_dept;
  vector[N_dept] beta_age_raw;
  real intercept_beta_age;
  real<lower=0> sigma_beta_age;
  real slope_beta_age;

  vector[N_dept] ihr_dept_raw;
  real intercept_ihr;
  real beta_ihr_age;
  real beta_ihr_beds;
  real beta_ihr_diabetes;
  real slope_ihr;
  real<lower=0> sigma_ihr_dept;

  vector[N_dept] ifr_dept_raw;
  real intercept_ifr;
  real beta_ifr_age;
  real beta_ifr_beds;
  real beta_ifr_diabetes;
  real slope_ifr;
  real<lower=0> sigma_ifr_dept;
```

```
}

transformed parameters {

  vector[N_dept] phi = sigma_phi * phi_raw;
  vector[N_dept] alpha_dept = mu_dept + phi;

  vector[N_dept] beta_age = beta_age_raw * sigma_beta_age +
  intercept_beta_age + slope_beta_age * alpha_dept;

  matrix<lower=0, upper=1>[N_age, N_dept] cuminc_age_dept;
  for (i in 1:N_age) {
    for (j in 1:N_dept) {
      cuminc_age_dept[i, j] = inv_logit(
        alpha_dept[j] + beta_age[j] * (i - 1)
      );
    }
  }

  matrix<lower=0, upper=1>[N_age, N_dept] seroprev_age_dept;
  for (i in 1:N_age) {
    for (j in 1:N_dept) {
      seroprev_age_dept[i, j] =
        sens * cuminc_age_dept[i, j] +
        (1 - spe) * (1 - cuminc_age_dept[i, j]);
    }
  }

  vector<lower=0, upper=1>[N_dept] seroprev_dept;
  for (i in 1:N_dept) {
    seroprev_dept[i] = mat_age_dept[, i]' * seroprev_age_dept[, i];
  }

  real seroprev_global_via_dept;
  seroprev_global_via_dept = seroprev_dept' * prop_dept;

  vector<lower=0, upper=1>[N_dept] cuminc_dept;
  for (i in 1:N_dept) {
    cuminc_dept[i] = mat_age_dept[, i]' * cuminc_age_dept[, i];
  }
```

```
  vector[N_dept] logit_ihr_dept;
  for (j in 1:N_dept) {
    logit_ihr_dept[j] =
      ihr_dept_raw[j] * sigma_ihr_dept +
      intercept_ihr +
      beta_ihr_age * age_pop[j] + beta_ihr_beds * beds[j] +
      beta_ihr_diabetes * diabetes[j] +
      slope_ihr * cuminc_dept[j];
  }

  vector[N_dept] logit_ifr_dept;
  for (j in 1:N_dept) {
    logit_ifr_dept[j] =
      ifr_dept_raw[j] * sigma_ifr_dept +
      intercept_ifr +
      beta_ifr_age * age_pop[j] + beta_ifr_beds * beds[j] +
      beta_ifr_diabetes * diabetes[j] +
      slope_ifr * cuminc_dept[j];
  }

  vector<lower=0, upper=1>[N_dept] ihr_dept = inv_logit(logit_ihr_dept) / 10;
  vector<lower=0, upper=1>[N_dept] ifr_dept = inv_logit(logit_ifr_dept) / 20;

}

model {

  phi_raw ~ icar_normal(N_dept, node1, node2);

  sens ~ beta(585, 56);
  spe ~ beta(953, 15);

  y_sens_sapris ~ binomial(N_sens_sapris, sens);

  seroprev_global_via_dept ~ beta(101, 1948);
  seroprev_global_via_dept ~ beta(1147, 17212);

  sigma_phi ~ exponential(1);
  sigma_ifr_dept ~ exponential(1);
  sigma_ihr_dept ~ exponential(1);
  beta_age_raw ~ normal(0, 1);
```

```
  for (k in 1:N_pos_incid) {
    1 ~ bernoulli(cuminc_age_dept[age_pos_incid[k] + 1, dept_pos_incid[k]]);
  }

  for (i in 1:N_age) {
    for (j in 1:N_dept) {
      mat_y_age_dept[i, j] ~ binomial(
        mat_N_age_dept[i, j], seroprev_age_dept[i, j]
      );
    }
  }

  ihr_dept_raw ~ normal(0, 1);
  ifr_dept_raw ~ normal(0, 1);

  for (j in 1:N_dept) {
    hospit_dept[j] ~ poisson(pop_dept[j] * cuminc_dept[j] * ihr_dept[j]);
    deces_dept[j] ~ poisson(pop_dept[j] * cuminc_dept[j] * ifr_dept[j]);
  }

}

generated quantities {

  // Association between incidence in persons under 60
  // and the proportion of persons above 60 among those infected

  vector<lower=0, upper=1>[N_dept] age_infected;
  for (j in 1:N_dept) {
    age_infected[j] =
      (cuminc_age_dept[2, j] * mat_age_dept[2, j]) /
      (cuminc_age_dept[2, j] * mat_age_dept[2, j] +
      cuminc_age_dept[1, j] * mat_age_dept[1, j]);
  }

  vector[N_grid_Y] g_cuminc_60_logit;
  for (i in 1:N_grid_Y) {
    g_cuminc_60_logit[i] =
      logit(grid_Y[i]) + intercept_beta_age +
      slope_beta_age * logit(grid_Y[i]);
  }
```

```
vector<lower=0, upper=1>[N_grid_Y] g_cuminc_60;
for (i in 1:N_grid_Y) {
  g_cuminc_60[i] = mean(inv_logit(normal_rng(
    rep_vector(g_cuminc_60_logit[i], N_draws),
    sigma_beta_age
  )));
}

vector<lower=0, upper=1>[N_grid_Y] g_age_infected;
for (i in 1:N_grid_Y) {
  g_age_infected[i] =
    (g_cuminc_60[i] * prop_age[2]) /
    (g_cuminc_60[i] * prop_age[2] + grid_Y[i] * prop_age[1]);
}

real douze_moins_six_age_infected = g_age_infected[41] - g_age_infected[17];

// Causal effects (incidence on IFR and IHR)

vector<lower=0, upper=1>[N_grid_Y] g_ihr_backdoor;
g_ihr_backdoor = rep_vector(0, N_grid_Y);

vector<lower=0, upper=1>[N_grid_Y] g_ifr_backdoor;
g_ifr_backdoor = rep_vector(0, N_grid_Y);

for (i in 1:N_dept) {
  for (s in 1:N_grid_Y) {

    real g_ihr_logit =
      intercept_ihr +
      beta_ihr_age * age_pop[i] + beta_ihr_beds * beds[i] +
      beta_ihr_diabetes * diabetes[i] +
      slope_ihr * grid_Y[s];

    real g_ifr_logit =
      intercept_ifr +
      beta_ifr_age * age_pop[i] + beta_ifr_beds * beds[i] +
      beta_ifr_diabetes * diabetes[i] +
      slope_ifr * grid_Y[s];

    real g_ihr = mean(inv_logit(to_vector(normal_rng(
```

```
      rep_vector(g_ihr_logit, N_draws), sigma_ihr_dept
    )))) / 10;

    real g_ifr = mean(inv_logit(to_vector(normal_rng(
      rep_vector(g_ifr_logit, N_draws), sigma_ifr_dept
    )))) / 20;

    g_ihr_backdoor[s] +=
      g_ihr / N_dept;

    g_ifr_backdoor[s] +=
      g_ifr / N_dept;

  }
}

real nine_minus_three_backdoor_ifr =
  g_ifr_backdoor[29] - g_ifr_backdoor[5];

real nine_minus_three_backdoor_ihr =
  g_ihr_backdoor[29] - g_ihr_backdoor[5];

// Global estimates for metropolitan France (incidence, IFR, IHR)

real cuminc_global_via_dept;
cuminc_global_via_dept = cuminc_dept' * prop_dept;

vector<lower=0>[N_dept] n_infected_dept;
for (j in 1:N_dept) {
  n_infected_dept[j] = cuminc_dept[j] * pop_dept[j];
}

real ifr_global;
ifr_global = (ifr_dept' * n_infected_dept) / sum(n_infected_dept);

real ihr_global;
ihr_global = (ihr_dept' * n_infected_dept) / sum(n_infected_dept);

// Posterior predictive samples
```

```
  array[N_age, N_dept] int<lower=0> y_rep_age_dept;
  for (i in 1:N_age) {
    for (j in 1:N_dept) {
      y_rep_age_dept[i ,j] = binomial_rng(
        mat_N_age_dept[i ,j], seroprev_age_dept[i ,j]
      );
    }
  }

  vector<lower=0>[N_dept] hospit_rep_dept;
  vector<lower=0>[N_dept] deces_rep_dept;

  for (j in 1:N_dept) {
    hospit_rep_dept[j] = poisson_rng(
      pop_dept[j] * cuminc_dept[j] * ihr_dept[j]
    );

    deces_rep_dept[j] = poisson_rng(
      pop_dept[j] * cuminc_dept[j] * ifr_dept[j]
    );
  }

}
```
